# Characterizing the Qatar advanced-phase SARS-CoV-2 epidemic

**DOI:** 10.1101/2020.07.16.20155317

**Authors:** Laith J. Abu Raddad, Hiam Chemaitelly, Houssein H. Ayoub, Zaina Al Kanaani, Abdullatif Al Khal, Einas Al Kuwari, Adeel A. Butt, Peter Coyle, Andrew Jeremijenko, Anvar Hassan Kaleeckal, Ali Nizar Latif, Robert C. Owen, Hanan F. Abdul Rahim, Samya A. Al Abdulla, Mohamed G. Al Kuwari, Mujeeb C. Kandy, Hatoun Saeb, Shazia Nadeem N. Ahmed, Hamad Eid Al Romaihi, Devendra Bansal, Louise Dalton, Sheikh Mohammad Al Thani, Roberto Bertollini

## Abstract

**Background:** Qatar has a population of 2.8 million, over half of whom are expatriate craft and manual workers (CMW). We aimed to characterize the severe acute respiratory syndrome coronavirus 2 (SARS-CoV-2) epidemic in Qatar.

**Methods:** A series of epidemiologic studies were conducted including analysis of the national SARS-CoV-2 PCR testing and hospitalization database, community surveys assessing current infection, ad-hoc PCR testing campaigns in workplaces and residential areas, serological testing for antibody on blood specimens collected for routine clinical screening/management, national Coronavirus Diseases 2019 (COVID-19) death registry, and a mathematical model.

**Results:** By July 10, 397,577 individuals had been PCR tested for SARS-CoV-2, of whom 110,986 were positive, a positivity cumulative rate of 27.9% (95% CI: 27.8-28.1%). PCR positivity of nasopharyngeal swabs in a national community survey (May 6-7) including 1,307 participants was 14.9% (95% CI: 11.5-19.0%); 58.5% of those testing positive were asymptomatic. Across 448 ad-hoc PCR testing campaigns in workplaces and residential areas including 26,715 individuals, pooled mean PCR positivity was 15.6% (95% CI: 13.7-17.7%). SARS-CoV-2 antibody prevalence was 24.0% (95% CI: 23.3-24.6%) in 32,970 residual clinical blood specimens. Antibody prevalence was only 47.3% (95% CI: 46.2-48.5%) in those who had at least one PCR positive result, but it was 91.3% (95% CI: 89.5-92.9%) among those who were PCR positive >3 weeks before serology testing. There were substantial differences in exposure to infection by nationality and sex, reflecting risk differentials between the craft/manual workers and urban populations. As of July 5, case severity rate, based on the WHO severity classification, was 3.4% and case fatality rate was 1.4 per 1,000 persons. Model-estimated daily number of infections and active-infection prevalence peaked at 22,630 and 5.7%, respectively, on May 21 and May 23. Attack rate (ever infection) was estimated at 53.5% on July 12. *R_0_* ranged between 1.45-1.68 throughout the epidemic. *R_t_* was estimated at 0.70 on June 15, which was hence set as onset date for easing of restrictions. Age was by far the strongest predictor of severe, critical, or fatal infection.

**Conclusions:** Qatar has experienced a large SARS-CoV-2 epidemic that is rapidly declining, apparently due to exhaustion of susceptibles. The epidemic demonstrated a classic susceptible-infected-recovered “SIR” dynamics with a rather stable *R_0_* of about 1.6. The young demographic structure of the population, in addition to a resourced public health response, yielded a milder disease burden and lower mortality than elsewhere.

## Introduction

Qatar is an Arabian Gulf country of 2.8 million people that has been affected by the severe acute respiratory syndrome coronavirus 2 (SARS-CoV-2) pandemic. The first documented case of SARS-CoV-2 community transmission was identified on March 6, 2020 and its source was linked soon after to a cluster of over 300 infections among expatriate craft and manual workers (CMW) living in high-density housing accommodations. Using mathematical modelling, the cluster size suggested the infection may have been circulating for at least 4 weeks prior to cluster identification. Restrictive social and physical distancing and other public health measures were immediately imposed in the whole country. By July 11, 103,128 SARS-CoV-2 infections had been laboratory-confirmed, at a rate of 36,729 per million population—one of the highest worldwide.^1^ Following the World Health Organization (WHO) guidelines, Qatar adopted a “testing, tracing, and isolation” approach, as the backbone of the national response,^2^ implementing a country-wide active contact tracing and testing using polymerase chain reaction (PCR). A total of 409,199 tests have been conducted for SARS-COV-2 using PCR as of July 11, at a rate of 145,736 per million population—one of the highest worldwide.^1^

Qatar has a unique demographic and residential dwellings structure^3^ that proved critical in understanding SARS-CoV-2 epidemiology. Of the total population,^3^ 89% are expatriates from over 150 countries,^4-6^ most of whom live in the capital city, Doha.^3^ About 60% of the population consists of CMW, typically working in mega-development projects.^7^ This “labor” population is predominantly young (20-49 years of age), male, and single, living generally in communal housing accommodations akin to dormitories.^8^ The remaining 40% of the population constitutes the “urban” population, which consists of family households or discrete-unit housings that include children, adults, and elderly, with adults often working in professional or service sector jobs, either private or governmental. Overall, including the CMW, Qatar’s population is predominantly young with only 2% being >60 years of age.^4^

Females account for only a quarter of the total population and for the vast majority are part of the urban population.^4^ By nationality, Indians (28%), Bangladeshis (13%), and Nepalese (13%) are the most populous groups followed by Qataris (11%), Egyptians (9%), and Filipinos (7%).^6^ Nearly all Bangladeshis and Nepalese are in the labor population. Meanwhile, Indians, Egyptians, and Filipinos are distributed among the labor and urban populations, but with most Indians in the labor population and most Filipinos in the urban population.^7^ Much of the remaining nationalities, who come from >150 countries,^4-6^ along with Qataris, are part of the urban population.^7^

In this article, we report the main findings of a series of epidemiologic studies to characterize SARS-CoV-2 epidemiology in Qatar. These studies include analyses of 1) the national SARS-CoV-2 PCR testing and hospitalization database, 2) community PCR testing surveys for current infection, 3) surveillance PCR testing campaigns in workplaces and residential areas, 4) serological testing for antibody on blood specimens collected for routine clinical screening/management, 5) national Coronavirus Disease 2019 (COVID-19) death registry, 6) SARS-CoV-2 case severity and mortality rates, and 7) a mathematical model investigating and forecasting the SARS-CoV-2 epidemic.

## Methods

### Sources of data

#### National SARS-CoV-2 PCR testing and hospitalization database

A national SARS-CoV-2 PCR testing and hospitalization database—a centralized and standardized database of all SARS-CoV-2 infections—is compiled at Hamad Medical Corporation (HMC), the main public healthcare provider and the nationally-designated provider for COVID-19 healthcare needs. The database comprises information on PCR testing conducted from February 5-July 10, 2020, including testing of suspected SARS-CoV-2 cases and traced contacts, in addition to infection surveillance testing. The database also includes data on hospital admission of COVID-19 patients and WHO severity classification for each infection.^9^

#### Community surveys

A cross-sectional community survey was conducted on May 6-7 to assess SARS-CoV-2 infection levels by PCR testing in the wider population of Qatar. Recruitment sampling frame was a database of 1,461,763 registered users (mostly covering the urban population) across Qatar’s 27 Primary Health Care Corporation (PHCC) centers. A sample size of 1,118 was needed to detect a population prevalence of 3% with a margin of error of 1%. To account for non-response, a phone-message invite was sent to 3,120 individuals. However, when only 268 invitees (24.0% of required sample size) responded to the invitation, recruitment was extended to the community by open invitation in national and social media.

Individuals were eligible to participate if they had not been previously diagnosed with SARS-CoV-2 infection, were 10-74 years of age, and registered users of PHCC. Demographic, exposure, and symptoms data and nasopharyngeal and oropharyngeal swabs were collected by trained personnel through a drive-through set-up in three PHCC centers located close to the Central, Northern, and Western regions of Qatar. Since the epidemic start, PHCC centers were designated as testing facilities for suspected cases. The design of the survey and interview schedule were informed by WHO guidelines^10^ and a SARS-CoV-2 population-based survey in Iceland^11^ (Text S1), with interviews administered in English or Arabic, depending on the respondent’s preference.

We further report the results of two other limited-scope community PCR testing surveys conducted among single CMW in a lockdown zone of Doha, where the first cluster of infections was identified. In the first one conducted between March 22-27, 5,120 individuals were sampled and tested using nasopharyngeal and oropharyngeal swabs. In a second survey in the same community on April 6-7, 886 individuals were sampled and tested. These two surveys were intended to be based on probability-based sampling, but logistical challenges forced a pragmatic, yet still heterogeneous by location, convenience sampling.

#### Ad-hoc testing campaigns in workplaces and in residential areas

A national SARS-CoV-2 testing database was compiled including the individual-level PCR testing outcomes of 390 ad-hoc testing campaigns of 22,834 individuals invited randomly to participate in a variety of workplaces in different economic sectors, and the outcomes of 58 ad-hoc testing campaigns covering 3,881 individuals invited randomly to participate in a variety of residential areas (often where CMW live). These testing campaigns were initiated by the Ministry of Public Health (MOPH) shortly after the identification of the first cluster in March, but accelerated in subsequent weeks. The available database covers all testing conducted up to June 4.

#### Seroprevalence survey

SARS-CoV-2 serological testing for antibody was performed on a convenience sample of residual blood specimens collected for routine clinical screening or clinical management from 32,970 outpatient and inpatient departments at HMC for a variety of health conditions, between May 12-July 12. Specimens were unlinked of identifying information about prior PCR testing before blood collection. The sample under-represented the CMW population as they receive outpatient healthcare primarily at customized healthcare centers operated by the Qatar Red Crescent Society. The database was subsequently linked to the national SARS-CoV-2 PCR testing and hospitalization database to conduct additional analyses linking PCR and antibody test results.

#### National COVID-19 mortality database

COVID-19 deaths were extracted from a national COVID-19 mortality validation database, a centralized registry of COVID-19-related deaths per WHO classification,^12^ compiled at HMC. The database comprises information on COVID-19 deaths from February 26-July 10, 2020.

### Laboratory methods

PCR testing was conducted at HMC Central Laboratory, the national reference laboratory, or at Sidra Medicine Laboratory, following standardized protocols. Nasopharyngeal and/or oropharyngeal swabs (Huachenyang Technology, China) were collected and placed in Universal Transport Medium (UTM). Aliquots of UTM were: extracted on the QIAsymphony platform (QIAGEN, USA) and tested with real-time reverse-transcription PCR (RT-qPCR) using the TaqPath(tm) COVID-19 Combo Kit (Thermo Fisher Scientific, USA) on a ABI 7500 FAST (ThermoFisher, USA); extracted using a custom protocol^13^ on a Hamilton Microlab STAR (Hamilton, USA) and tested using the AccuPower SARS-CoV-2 Real-Time RT-PCR Kit (Bioneer, Korea) on a ABI 7500 FAST; or loaded directly to a Roche cobas® 6800 system and assayed with the cobas® SARS-CoV-2 Test (Roche, Switzerland). The first assay targets the S, N, and ORF1ab regions of the virus; the second targets the virus’ RdRp and E-gene regions; and the third targets the ORF1ab and E-gene regions.

Serological testing was performed using the Roche Elecsys^®^ Anti-SARS-CoV-2 (Roche, Switzerland), an electrochemiluminescence immunoassay that uses a recombinant protein representing the nucleocapsid (N) antigen for the determination of antibodies against SARS-CoV-2. Qualitative anti-SARS-CoV-2 results were generated following the manufacturer’s instructions^14^ (reactive: cutoff index ≥1.0 vs. non-reactive: cutoff index <1.0).

### Statistical analysis

Frequency distributions were generated to describe the demographic/clinical profile of tested individuals. Where applicable, probability weights were applied to adjust for unequal sample selection using the Qatar population distribution by sex, age group, and nationality.^4,6,15^ Chi-square test and univariable logistic regressions were implemented to explore associations. Odds ratios (ORs), 95% confidence intervals (CIs), and p-values were reported. Covariates with p-value ≤0.1 in univariable regression analysis were considered possibly associated with the outcome variables, and were thus included in the multivariable analysis for estimation of adjusted odds ratios (AORs) and associated 95% CIs and p-values. Covariates with p-value ≤0.05 in the multivariable model were considered as predictors of the outcome.

Where relevant, the pooled mean for SARS-CoV-2 PCR positivity was estimated using random-effects meta-analysis. To this end, variances of measures were first stabilized using a Freeman-Tukey type arcsine square-root transformation.^16,17^ Measures were then weighted using the inverse-variance method,^17,18^ prior to being pooled using a DerSimonian-Laird random-effects model.^19^ Factors associated with higher PCR positivity and sources of between-study heterogeneity were then identified using random-effects meta-regression, applying the same criteria used for conventional regression analysis (described above).

### Mathematical model

An age-structured deterministic mathematical model was constructed to describe the time-course of the SARS-CoV-2 epidemic. Susceptible individuals in each age group were at risk of acquiring the infection based on their infectious contact rate per day, age-specific susceptibility to the infection, and an age-mixing matrix defining mixing between individuals in the different age groups (Text S2). Following a latency period, infected individuals develop asymptomatic/mild, severe, or critical infection. Severe and critical infections progress to severe and critical disease, respectively, prior to recovery; these are hospitalized, in acute and intensive care unit (ICU) care beds, respectively. Critical disease cases have an additional risk for COVID-19 mortality. Population movement between model compartments was described using a set of coupled nonlinear differential equations (Text S2).

The model was parameterized using available data for SARS-CoV-2 natural history and epidemiology. Description of model parameters, definitions, and values are in Text S3 and Tables S1-S2.

### Ethical approval

Studies were approved by HMC and Weill Cornell Medicine-Qatar Institutional Review Boards.

## Results

### Analysis of the national SARS-CoV-2 PCR testing and hospitalization database

By July 10, a total of 397,577 individuals had been tested for current infection (14.2% of the population of Qatar), of whom 110,986 were PCR positive for SARS-CoV-2 (4.0% of the population), for an overall cumulative positivity rate of 27.9% (95% CI: 27.8-28.1%). Positivity rate increased rapidly starting from March and peaked at 45.1% on May 22, after which it has been declining and was 17.8% on July 9.

Adjusted odds of PCR positivity were 1.6-fold (95% CI: 1.5-1.6) higher in males compared to females (Table 1). Odds of PCR positivity varied by nationality and were highest among Nepalese (AOR: 4.5; 95% CI: 4.3-4.6) and Bangladeshis (AOR: 3.9; 95% CI: 3.8-4.0) and lowest among Qataris (AOR: 0.57; 95% CI: 0.55-0.59), compared to other nationalities.

**Table 1.**
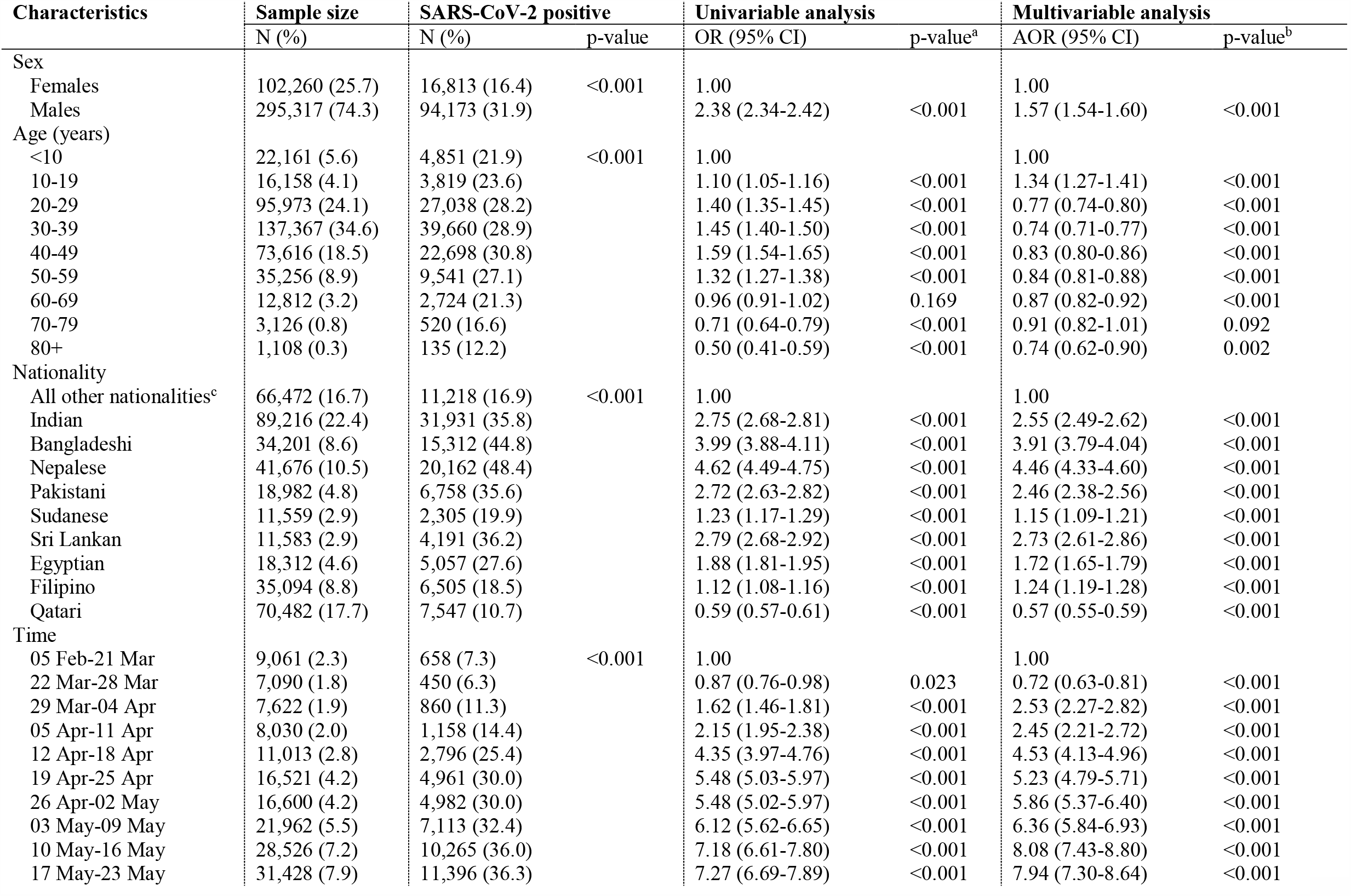
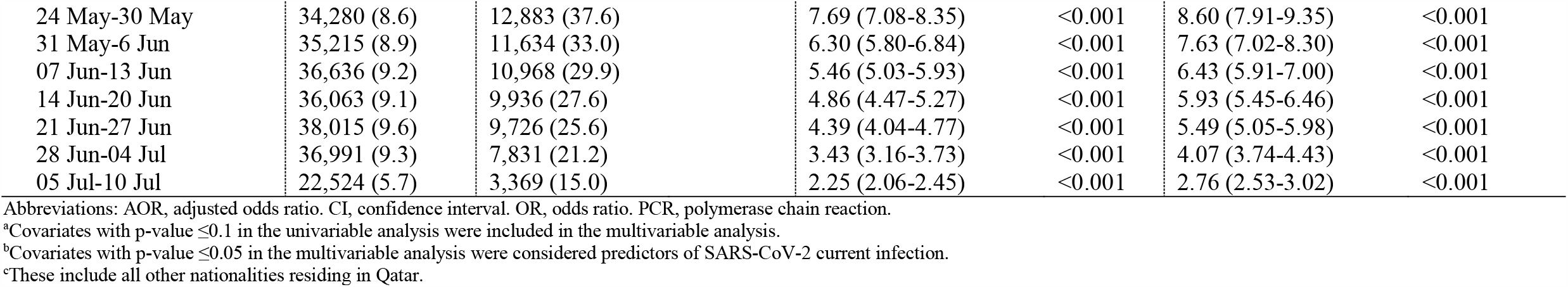
Associations with current infection in Qatar based on analysis of the national SARS-CoV-2 PCR testing and hospitalization database

| Characteristics | Sample size | SARS-CoV-2 positive |  | Univariable analysis |  | Multivariable analysis |  |
| --- | --- | --- | --- | --- | --- | --- | --- |
|  | N (%) | N (%) | p-value | OR (95% CI) | p-value<sup>a</sup> | AOR (95% CI) | p-value<sup>b</sup> |
| Sex |  |  |  |  |  |  |  |
| Females | 102,260 (25.7) | 16,813 (16.4) | <0.001 | 1.00 |  | 1.00 |  |
| Males | 295,317 (74.3) | 94,173 (31.9) |  | 2.38 (2.34-2.42) | <0.001 | 1.57 (1.54-1.60) | <0.001 |
| Age (years) |  |  |  |  |  |  |  |
| <10 | 22,161 (5.6) | 4,851 (21.9) | <0.001 | 1.00 |  | 1.00 |  |
| 10-19 | 16,158 (4.1) | 3,819 (23.6) |  | 1.10 (1.05-1.16) | <0.001 | 1.34 (1.27-1.41) | <0.001 |
| 20-29 | 95,973 (24.1) | 27,038 (28.2) |  | 1.40 (1.35-1.45) | <0.001 | 0.77 (0.74-0.80) | <0.001 |
| 30-39 | 137,367 (34.6) | 39,660 (28.9) |  | 1.45 (1.40-1.50) | <0.001 | 0.74 (0.71-0.77) | <0.001 |
| 40-49 | 73,616 (18.5) | 22,698 (30.8) |  | 1.59 (1.54-1.65) | <0.001 | 0.83 (0.80-0.86) | <0.001 |
| 50-59 | 35,256 (8.9) | 9,541 (27.1) |  | 1.32 (1.27-1.38) | <0.001 | 0.84 (0.81-0.88) | <0.001 |
| 60-69 | 12,812 (3.2) | 2,724 (21.3) |  | 0.96 (0.91-1.02) | 0.169 | 0.87 (0.82-0.92) | <0.001 |
| 70-79 | 3,126 (0.8) | 520 (16.6) |  | 0.71 (0.64-0.79) | <0.001 | 0.91 (0.82-1.01) | 0.092 |
| 80+ | 1,108 (0.3) | 135 (12.2) |  | 0.50 (0.41-0.59) | <0.001 | 0.74 (0.62-0.90) | 0.002 |
| Nationality |  |  |  |  |  |  |  |
| All other nationalities<sup>c</sup> | 66,472 (16.7) | 11,218 (16.9) | <0.001 | 1.00 |  | 1.00 |  |
| Indian | 89,216 (22.4) | 31,931 (35.8) |  | 2.75 (2.68-2.81) | <0.001 | 2.55 (2.49-2.62) | <0.001 |
| Bangladeshi | 34,201 (8.6) | 15,312 (44.8) |  | 3.99 (3.88-4.11) | <0.001 | 3.91 (3.79-4.04) | <0.001 |
| Nepalese | 41,676 (10.5) | 20,162 (48.4) |  | 4.62 (4.49-4.75) | <0.001 | 4.46 (4.33-4.60) | <0.001 |
| Pakistani | 18,982 (4.8) | 6,758 (35.6) |  | 2.72 (2.63-2.82) | <0.001 | 2.46 (2.38-2.56) | <0.001 |
| Sudanese | 11,559 (2.9) | 2,305 (19.9) |  | 1.23 (1.17-1.29) | <0.001 | 1.15 (1.09-1.21) | <0.001 |
| Sri Lankan | 11,583 (2.9) | 4,191 (36.2) |  | 2.79 (2.68-2.92) | <0.001 | 2.73 (2.61-2.86) | <0.001 |
| Egyptian | 18,312 (4.6) | 5,057 (27.6) |  | 1.88 (1.81-1.95) | <0.001 | 1.72 (1.65-1.79) | <0.001 |
| Filipino | 35,094 (8.8) | 6,505 (18.5) |  | 1.12 (1.08-1.16) | <0.001 | 1.24 (1.19-1.28) | <0.001 |
| Qatari | 70,482 (17.7) | 7,547 (10.7) |  | 0.59 (0.57-0.61) | <0.001 | 0.57 (0.55-0.59) | <0.001 |
| Time |  |  |  |  |  |  |  |
| 05 Feb-21 Mar | 9,061 (2.3) | 658 (7.3) | <0.001 | 1.00 |  | 1.00 |  |
| 22 Mar-28 Mar | 7,090 (1.8) | 450 (6.3) |  | 0.87 (0.76-0.98) | 0.023 | 0.72 (0.63-0.81) | <0.001 |
| 29 Mar-04 Apr | 7,622 (1.9) | 860 (11.3) |  | 1.62 (1.46-1.81) | <0.001 | 2.53 (2.27-2.82) | <0.001 |
| 05 Apr-11 Apr | 8,030 (2.0) | 1,158 (14.4) |  | 2.15 (1.95-2.38) | <0.001 | 2.45 (2.21-2.72) | <0.001 |
| 12 Apr-18 Apr | 11,013 (2.8) | 2,796 (25.4) |  | 4.35 (3.97-4.76) | <0.001 | 4.53 (4.13-4.96) | <0.001 |
| 19 Apr-25 Apr | 16,521 (4.2) | 4,961 (30.0) |  | 5.48 (5.03-5.97) | <0.001 | 5.23 (4.79-5.71) | <0.001 |
| 26 Apr-02 May | 16,600 (4.2) | 4,982 (30.0) |  | 5.48 (5.02-5.97) | <0.001 | 5.86 (5.37-6.40) | <0.001 |
| 03 May-09 May | 21,962 (5.5) | 7,113 (32.4) |  | 6.12 (5.62-6.65) | <0.001 | 6.36 (5.84-6.93) | <0.001 |
| 10 May-16 May | 28,526 (7.2) | 10,265 (36.0) |  | 7.18 (6.61-7.80) | <0.001 | 8.08 (7.43-8.80) | <0.001 |
| 17 May-23 May | 31,428 (7.9) | 11,396 (36.3) |  | 7.27 (6.69-7.89) | <0.001 | 7.94 (7.30-8.64) | <0.001 |

|  |  |  |  |  |  |  |
| --- | --- | --- | --- | --- | --- | --- |
| 24 May-30 May | 34,280 (8.6) | 12,883 (37.6) | 7.69 (7.08-8.35) | <0.001 | 8.60 (7.91-9.35) | <0.001 |
| 31 May-6 Jun | 35,215 (8.9) | 11,634 (33.0) | 6.30 (5.80-6.84) | <0.001 | 7.63 (7.02-8.30) | <0.001 |
| 07 Jun-13 Jun | 36,636 (9.2) | 10,968 (29.9) | 5.46 (5.03-5.93) | <0.001 | 6.43 (5.91-7.00) | <0.001 |
| 14 Jun-20 Jun | 36,063 (9.1) | 9,936 (27.6) | 4.86 (4.47-5.27) | <0.001 | 5.93 (5.45-6.46) | <0.001 |
| 21 Jun-27 Jun | 38,015 (9.6) | 9,726 (25.6) | 4.39 (4.04-4.77) | <0.001 | 5.49 (5.05-5.98) | <0.001 |
| 28 Jun-04 Jul | 36,991 (9.3) | 7,831 (21.2) | 3.43 (3.16-3.73) | <0.001 | 4.07 (3.74-4.43) | <0.001 |
| 05 Jul-10 Jul | 22,524 (5.7) | 3,369 (15.0) | 2.25 (2.06-2.45) | <0.001 | 2.76 (2.53-3.02) | <0.001 |

Abbreviations: AOR, adjusted odds ratio. CI, confidence interval. OR, odds ratio. PCR, polymerase chain reaction.
<sup>a</sup>Covariates with p-value $\leq 0.1$ in the univariable analysis were included in the multivariable analysis.
<sup>b</sup>Covariates with p-value $\leq 0.05$ in the multivariable analysis were considered predictors of SARS-CoV-2 current infection.
<sup>c</sup>These include all other nationalities residing in Qatar.

A time trend was observed with odds of PCR positivity gradually increasing (Table 1), consistent with an exponentially growing epidemic, to reach 8.6-fold (95% CI: 7.9-9.4) higher in May 24-30 compared to the beginning of the epidemic, but rapidly declining thereafter to be only 2.8-fold (95% CI: 2.5-3.0) higher in July 05-10.

### PCR positivity in community surveys

A total of 1,307 individuals participated in the PCR community survey conducted on May 6-7 (Table 2). There were differences in age, nationality, and educational attainment between the participants who were randomly invited and those who participated through the open announcement, but the differences were not major (Table S3). No differences were observed by sex or occupation.

**Table 2.**
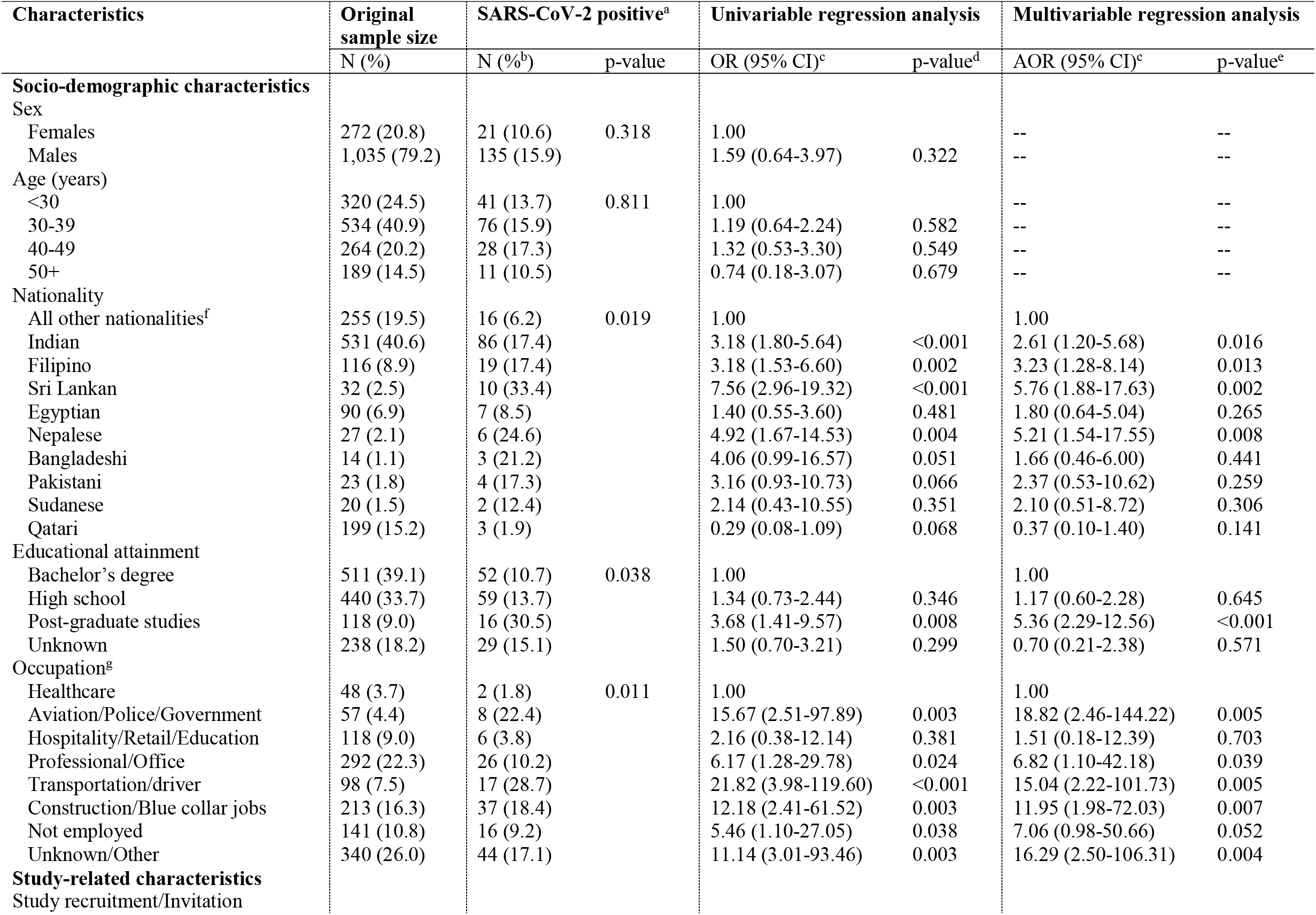
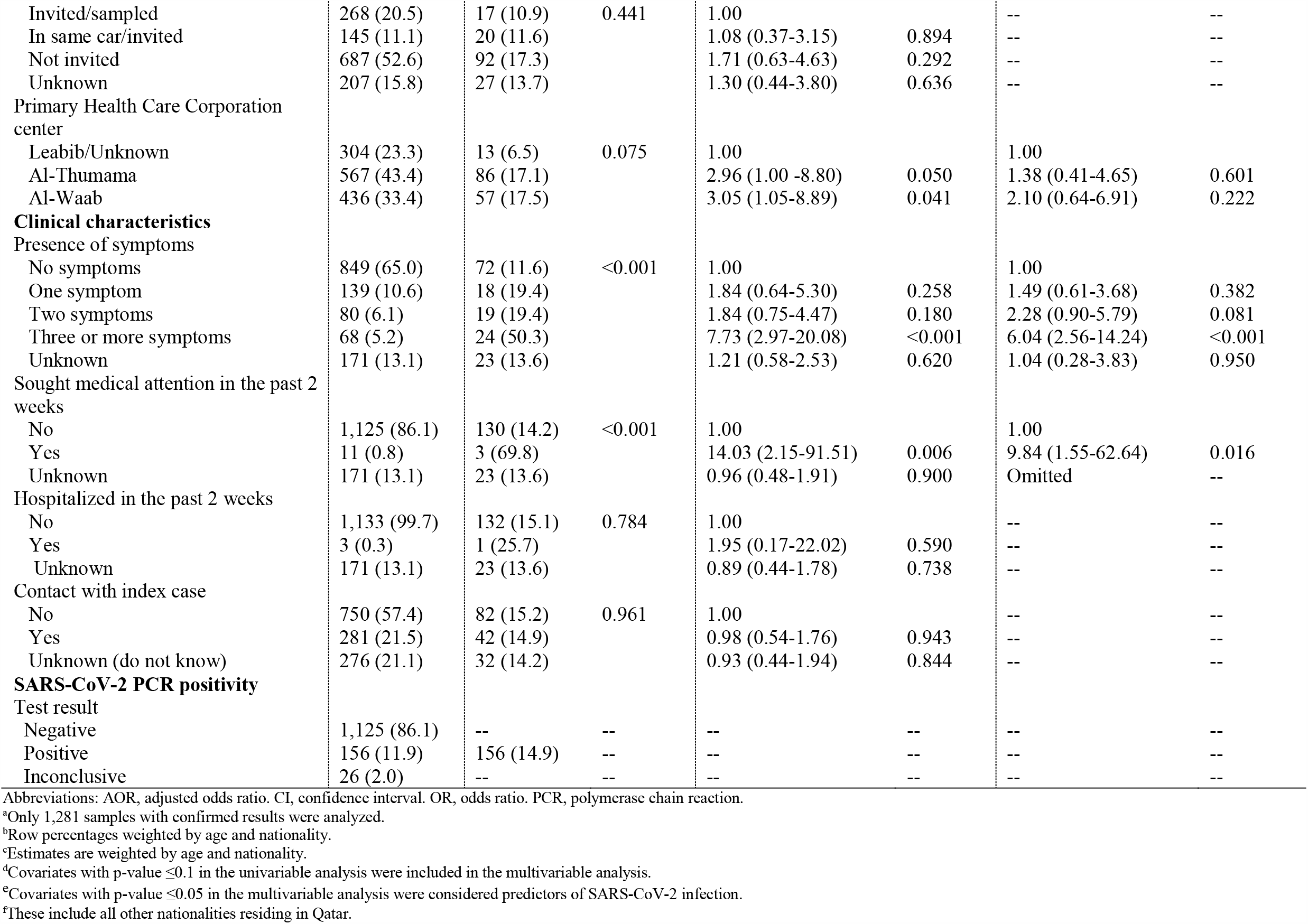
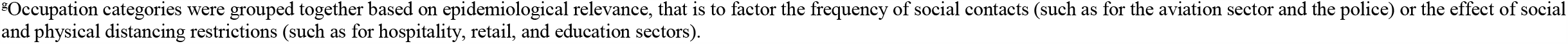
Results of the community survey conducted on May 6-7, 2020 and associations with PCR positivity in Qatar

A total of 156 persons tested PCR positive. The weighted (for total population of Qatar) SARS-CoV-2 PCR prevalence was 14.9% (95% CI: 11.5-19.0%). After controlling for confounders, strong evidence for an association with positivity (p-value ≤0.05) was found for nationality, educational attainment, occupation, presence of symptoms, and seeking medical attention, but not for sex, age, study recruitment/invitation, PHCC center location, seeking hospitalization, or history of contact with an index case (Table 2). The differences were pronounced for nationality, occupation, presence of symptoms, and seeking medical attention.

The proportion of asymptomatic individuals was 58.5% in those testing positive and 79.5% in those testing negative (p<0.001; Table 3). While there was no evidence for an association with reporting one or two symptoms, reporting three or more symptoms was significantly associated with SARS-CoV-2 infection (Table 3). Associations with specific symptoms are summarized in Table S4—infection was significantly associated with fever, fatigue, muscle ache, other respiratory symptoms, nausea/vomiting, loss of sense of smell, and loss of sense of taste.

**Table 3.** Comparison of individuals testing PCR negative versus positive for SARS-CoV-2^a^ in the community survey conducted on May 6-7, 2020 with respect to presence of symptoms

| Reported symptoms | SARS-CoV-2<br>negative | SARS-CoV-2<br>positive | p-value | Univariable regression<br>analysis |  |
| --- | --- | --- | --- | --- | --- |
|  | N (%<sup>b</sup>) | N (%<sup>b</sup>) |  | OR (95% CI)<sup>c</sup> | p-value |
| Presence of symptoms |  |  |  |  |  |
| Asymptomatic | 764 (79.5) | 72 (58.5) | 0.003 | 1.00 |  |
| Symptomatic | 217 (20.5) | 61 (41.6) |  | 2.75 (1.39-5.45) | 0.004 |
| Overall |  |  |  |  |  |
| No symptoms | 764 (79.5) | 72 (58.5) | <0.001 | 1.00 |  |
| One symptom | 119 (11.0) | 18 (14.9) |  | 1.84 (0.64-5.30) | 0.258 |
| Two symptoms | 59 (6.3) | 19 (8.5) |  | 1.84 (0.75-4.47) | 0.180 |
| Three or more symptoms | 39 (3.2) | 24 (18.1) |  | 7.73 (2.97-20.08) | <0.001 |

Abbreviations: CI, confidence interval. OR, odds ratio. PCR, polymerase chain reaction.
<sup>a</sup>Only 1,114 samples with information on symptoms and confirmed results were analyzed.
<sup>b</sup>Column percentages weighted by age and nationality.
<sup>c</sup>Estimates are weighted by age and nationality.

However, <9.5% of those positive reported each of these individual symptoms apart from fever which was reported by 28.6% of those positive.

An analysis of PCR cycle threshold (Ct) values’ association with presence of symptoms was performed with the understanding that PCR positivity does not only reflect recent active infection, but also time after recovery, as PCR tests are sensitive enough to pick even non-viable viral fragments even for weeks following recovery from active infection.^20,21^ Table S5 shows the association between PCR Ct value and presence of symptoms. Odds of having a Ct value <25 (proxy for recent infection^22^) were five-fold higher among symptomatic compared to asymptomatic individuals.

In the two other (limited-scope) community surveys conducted among single CMW in a lockdown zone of Doha, where the first cluster of infections was identified, PCR positivity was first assessed at 1.4% (95% CI: 1.1-1.8%) on March 22-27 in a sample comprising 5,120 individuals, and at 4.4% (95% CI: 3.2-6.0%) on April 6-7 in a sample comprising 886 individuals.

### PCR positivity in the ad-hoc testing campaigns in workplaces and in residential areas

By June 4, 390 random PCR testing campaigns in workplaces and 58 in residential areas had been conducted testing 22,834 and 3,881 individuals, respectively. At least one infected individual was identified in 72.3% of workplaces and 72.4% of residential areas. The median PCR point prevalence in workplaces was 7.8%, while the pooled mean was 15.1% (95% CI: 13.0-17.3%). Meanwhile, the median PCR point prevalence in residential areas was 15.3%, and the pooled mean was 20.3% (95% CI: 15.4-25.6%).

Table S6 shows the pooled mean PCR point prevalence across these testing campaigns and over time. The pooled mean was 17.6% (95% CI: 9.3-27.7%) in March, but testing campaigns then were focused in the specific neighborhood where the first cluster was identified. Starting from April, testing was expanded throughout Qatar. PCR point prevalence increased steadily and rapidly consistent with exponential growth from about 5% in early April to a peak of 23% by mid to late May, after which the prevalence started declining.

Table 4 shows the results of the multivariable meta-regression analysis of PCR point prevalence across these testing campaigns. There was no evidence for differences in PCR point prevalence between the workplaces and residential areas, consistent with a widely disseminated epidemic. However, there was strong evidence for a rapidly growing epidemic from March up to the third week of May, after which the epidemic started declining.

**Table 4.**
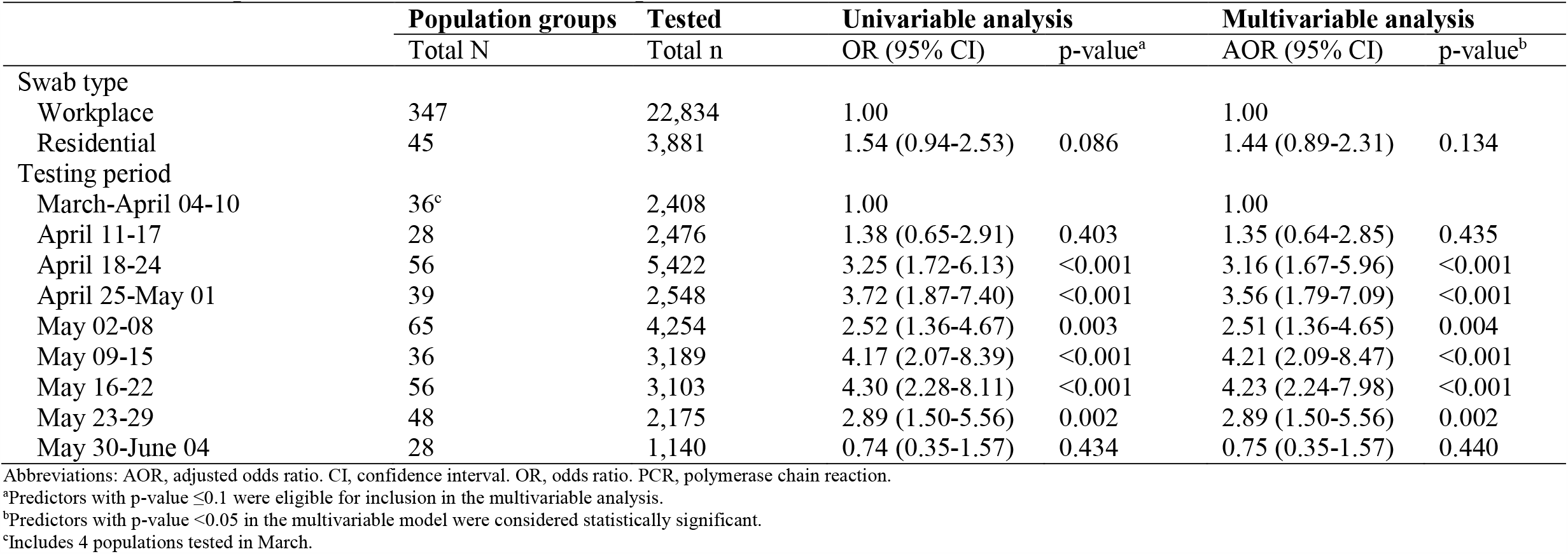
Meta-regression results to identify associations with SARS-CoV-2 PCR prevalence in the random testing campaigns conducted in workplaces and in residential areas, up to June 4, 2020

### Anti-SARS-COV-2 seropositivity

A total of 32,970 individuals were tested for SARS-CoV-2 antibodies with the blood specimens drawn between May 12-July 12, with a median day of June 28. A total of 5,448 individuals had detectable antibodies. The weighted (for total population of Qatar) antibody prevalence was 24.0% (95% CI: 23.3-24.6%).

There were large differences in antibody prevalence by sex and nationality. After controlling for confounders (Table 5), adjusted odds of antibody positivity were 2.9-fold (95% CI: 2.6-3.2) higher in males compared to females, and were highest among Bangladeshis (AOR: 6.8; 95% CI: 5.9-8.0) and Nepalese (AOR: 6.6; 95% CI: 5.7-7.8), but lowest among Qataris (AOR: 0.7; 95% CI: 0.6-0.8), compared to other nationalities. Adjusted odds of antibody positivity increased incrementally with age.

**Table 5.** Results of the seroprevalence survey and associations with antibody positivity

| Characteristics | Tested | Seropositive |  | Univariable regression analysis |  | Multivariable regression analysis |  |
| --- | --- | --- | --- | --- | --- | --- | --- |
|  | N (%) | N (%<sup>a</sup>) | p-value | OR<sup>a</sup> (95% CI) | p-value<sup>b</sup> | AOR<sup>a</sup> (95% CI) | p-value<sup>c</sup> |
| Sex |  |  |  |  |  |  |  |
| Females | 14,366 (43.6) | 958 (7.8) | <0.001 | 1.00 |  | 1.00 |  |
| Males | 18,604 (56.4) | 4,490 (29.2) |  | 4.89 (4.46-5.37) | <0.001 | 2.92 (2.64-3.24) | <0.001 |
| Age (years) |  |  |  |  |  |  |  |
| <10 | 814 (2.5) | 40 (5.5) | <0.001 | 1.00 |  | 1.00 |  |
| 10-19 | 1,222 (3.7) | 70 (6.7) |  | 1.25 (0.78-1.99) | 0.356 | 1.80 (1.11-2.93) | 0.018 |
| 20-29 | 4,738 (14.4) | 613 (23.1) |  | 5.22 (3.59-7.59) | <0.001 | 3.35 (2.26-4.96) | <0.001 |
| 30-39 | 8,437 (25.6) | 1,379 (25.3) |  | 5.87 (4.06-8.47) | <0.001 | 3.31 (2.25-4.87) | <0.001 |
| 40-49 | 7,513 (22.8) | 1,582 (30.1) |  | 7.47 (5.18-10.79) | <0.001 | 3.82 (2.60-5.61) | <0.001 |
| 50-59 | 5,455 (16.5) | 1,150 (30.4) |  | 7.57 (5.24-10.95) | <0.001 | 4.27 (2.91-6.28) | <0.001 |
| 60-69 | 2,982 (9.0) | 452 (23.9) |  | 5.46 (3.74-7.97) | <0.001 | 3.86 (2.60-5.74) | <0.001 |
| 70-79 | 1,254 (3.8) | 122 (13.9) |  | 2.79 (1.83-4.25) | <0.001 | 3.60 (2.33-5.56) | <0.001 |
| 80+ | 555 (1.7) | 40 (9.8) |  | 1.88 (1.13-3.12) | 0.015 | 3.36 (1.97-5.72) | <0.001 |
| Nationality |  |  |  |  |  |  |  |
| All other nationalities<sup>d</sup> | 6,776 (20.6) | 479 (7.2) | <0.001 | 1.00 |  | 1.00 |  |
| Indian | 5,553 (16.8) | 1,330 (24.3) |  | 4.16 (3.64-4.75) | <0.001 | 3.17 (2.76-3.63) | <0.001 |
| Bangladeshi | 2,284 (6.9) | 996 (44.6) |  | 10.44 (9.03-12.09) | <0.001 | 6.83 (5.86-7.96) | <0.001 |
| Nepalese | 1,622 (4.9) | 732 (44.1) |  | 10.22 (8.76-11.92) | <0.001 | 6.63 (5.65-7.77) | <0.001 |
| Pakistani | 1,524 (4.6) | 412 (23.8) |  | 4.04 (3.36-4.85) | <0.001 | 3.91 (3.24-4.73) | <0.001 |
| Sudanese | 1,123 (3.4) | 131 (11.0) |  | 1.60 (1.23-2.08) | 0.001 | 1.54 (1.18-2.02) | 0.002 |
| Sri Lankan | 696 (2.1) | 185 (26.5) |  | 4.68 (3.74-5.87) | <0.001 | 3.38 (2.70-4.23) | <0.001 |
| Egyptian | 2,612 (7.9) | 306 (11.7) |  | 1.72 (1.44-2.07) | <0.001 | 1.54 (1.28-1.85) | <0.001 |
| Filipino | 2,224 (6.7) | 465 (20.3) |  | 3.31 (2.81-3.89) | <0.001 | 3.31 (2.80-3.91) | <0.001 |
| Qatari | 8,556 (26.0) | 412 (4.7) |  | 0.63 (0.53-0.75) | <0.001 | 0.66 (0.55-0.79) | <0.001 |
| Antibody testing period |  |  |  |  |  |  |  |
| 12 May-31 May | 937 (2.8) | 264 (30.4) | <0.001 | 1.00 |  | 1.00 |  |
| 01 Jun-15 Jun | 3,485 (10.6) | 793 (30.2) |  | 0.99 (0.81-1.21) | 0.931 | 1.34 (1.08-1.67) | 0.009 |
| 16 Jun-30 Jun | 14,954 (45.4) | 2,261 (22.6) |  | 0.67 (0.56-0.80) | <0.001 | 1.05 (0.86-1.28) | 0.651 |
| 01 Jul-12 Jul | 13,594 (41.2) | 2,130 (23.1) |  | 0.69 (0.57-0.83) | <0.001 | 1.11 (0.91-1.35) | 0.316 |

Abbreviations: AOR, adjusted odds ratio. CI, confidence interval. OR, odds ratio.
<sup>a</sup>Estimates are weighted by sex, age, and nationality.<sup>b</sup>Covariates with p-value ≤0.1 in the univariable analysis were included in the multivariable analysis.<sup>c</sup>Covariates with p-value ≤0.05 in the multivariable analysis were considered predictors of anti-SARS-CoV-2 positivity.<sup>d</sup>These include all other nationalities residing in Qatar.

Linking the antibody testing database to the national SARS-CoV-2 PCR testing and hospitalization database identified 17,062 individuals with available PCR and antibody testing results. Antibody prevalence was 6.2% (95% CI: 5.7-6.7%) in those who had all PCR negative results. Antibody prevalence was 47.3% (95% CI: 46.2-48.5%) in those who had at least one PCR positive result.

Figure 1 shows antibody prevalence versus the time difference between the first positive PCR test and the antibody test. Overall, antibody positivity was >88% among those who had their first positive PCR test >15 days before the antibody test. Antibody positivity declined steadily the closer is the PCR test date to the antibody test date, consistent with a weeks-long delay between onset of infection and development of detectable antibodies.

**Figure 1.**
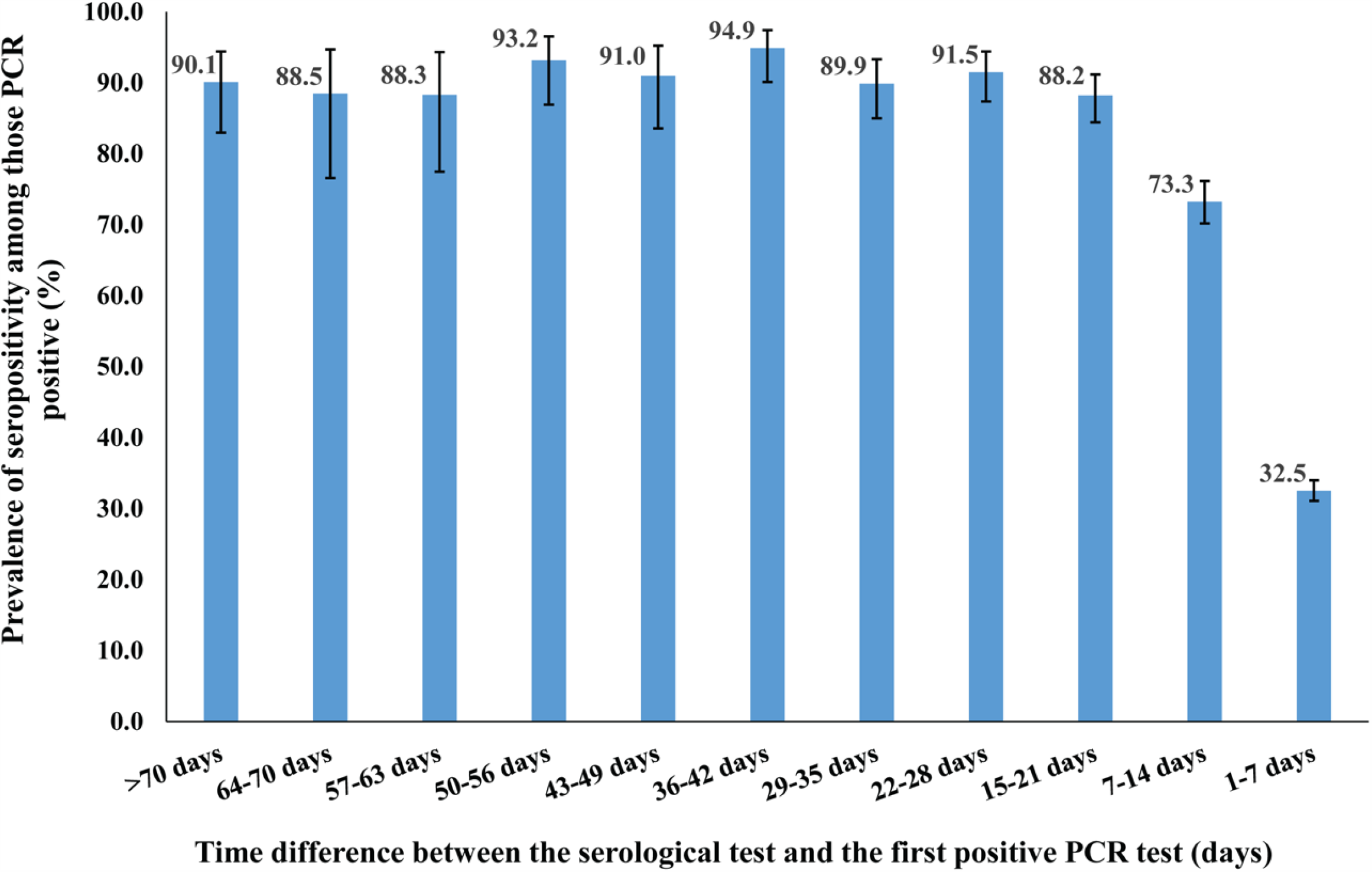
Anti-SARS-CoV-2 prevalence assessed at different time intervals for the duration between the first PCR positive test and the antibody test.

### Basic socio-demographic associations with severe, critical, or fatal infection

Tables S7-S9 show basic socio-demographic associations with each of severe and critical infection and COVID-19 death (all per WHO classification),^9,12^ respectively, as of June 25. After controlling for confounders, age was, by far, the strongest predictor of each of these outcomes, and more so for critical infection and death (Figure 2). Risk of serious disease and death increased immensely for those >50 years of age, who are a small minority in the population of Qatar (8.8%).

**Figure 2.**
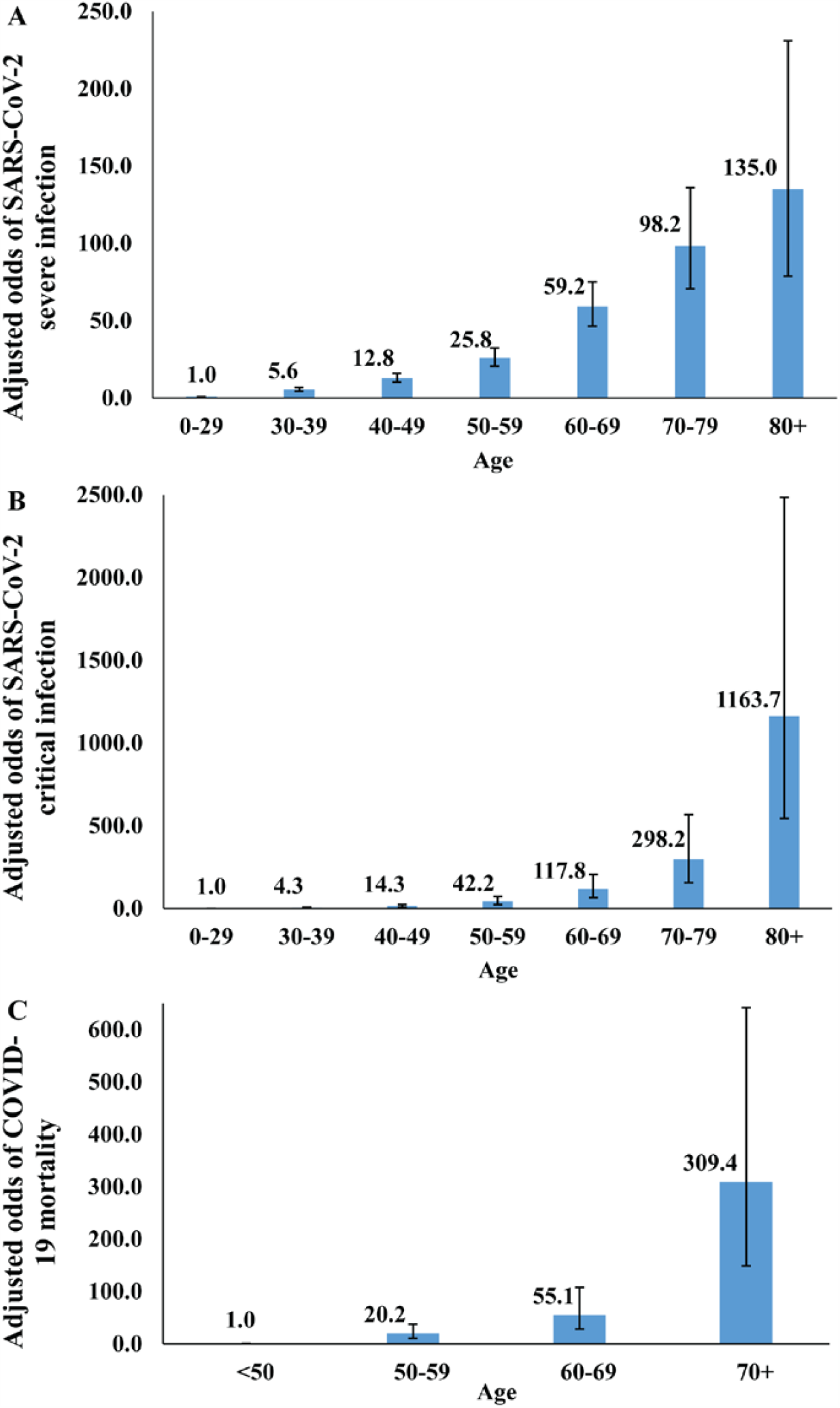
Association of age with A) SARS-CoV-2 severe infection, B) SARS-CoV-2 critical infection, and C) COVID-19 death, adjusted for sex, nationality, and PCR testing period.

Males had 1.6-fold higher odds for developing severe and critical infection compared to females (Tables S7-S8), but no difference was observed for death, possibly because of the small number of deaths (Table S9). Overall, there were no major differences in these outcomes by nationality, but there was evidence for higher risk of serious disease for Bangladeshis and Filipinos, and lower risk for Indians. The odds of disease and mortality declined with testing period, but this may just reflect the lagging time between onset of infection and disease/mortality.

### Crude case severity and fatality rates

Figure 3A shows the crude case severity rate versus time defined as the cumulative number of severe and critical infections over the cumulative number of laboratory-confirmed infections. The crude case severity rate was mostly stable, assessed at 3.4% on July 5.

**Figure 3.**
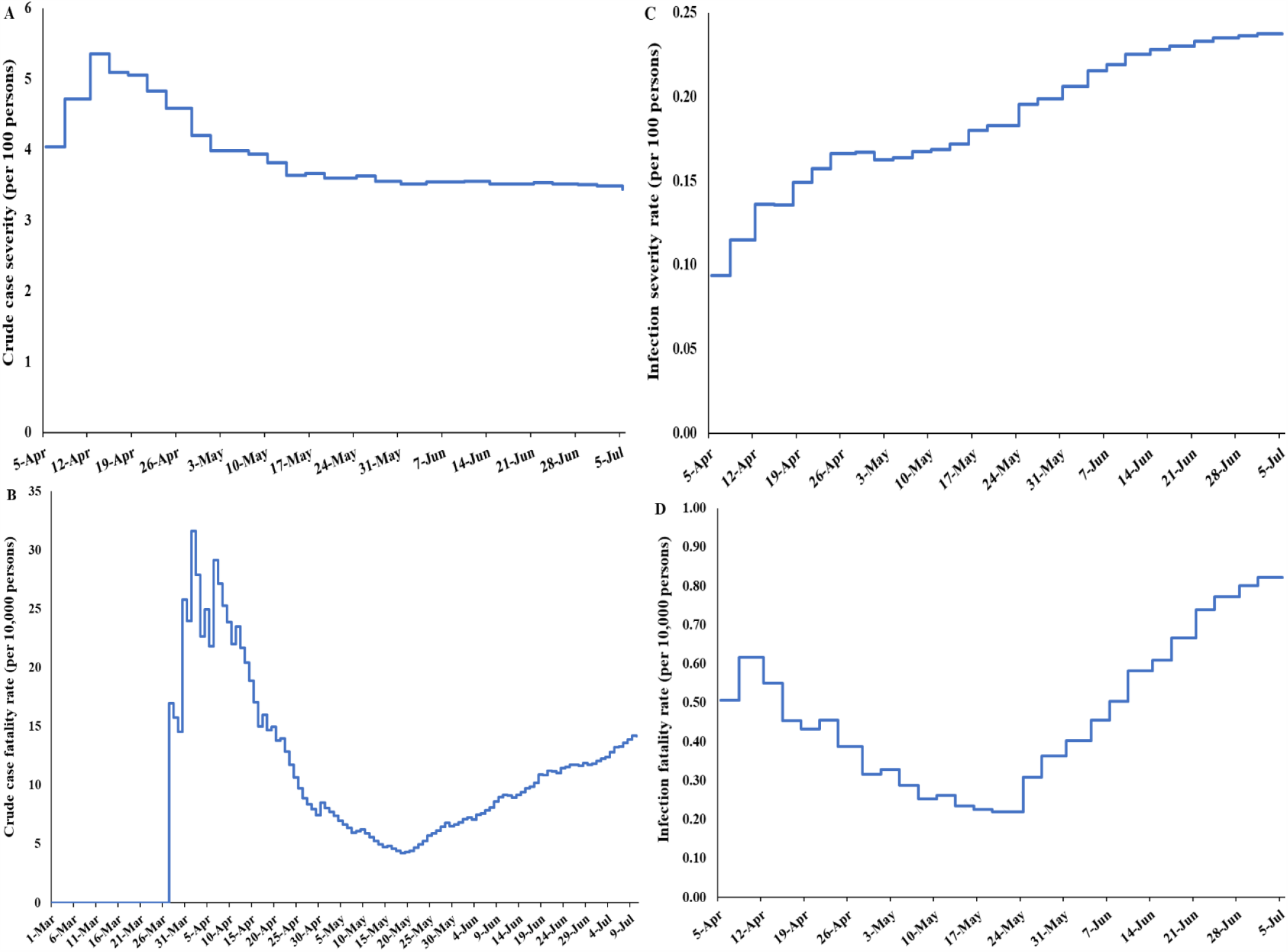
Temporal trend in A) crude case severity rate, B) crude case fatality rate, C) infection severity rate, and D) infection fatality rate in Qatar.

Figures 3B shows the crude case fatality rate versus time defined as the cumulative number of COVID-19 deaths over the cumulative number of laboratory-confirmed infections. The crude case fatality rate varied throughout the epidemic, in part because of the age structure of the population affected at each time point, but mostly because of statistical volatility with the relatively small number of COVID-19 deaths (146 as of July 10). The crude case fatality rate has been increasing steadily but slowly in recent weeks, consistent with the epidemic dynamics moving from the labor to the urban population where virtually all the elderly population resides. As of July 10, the crude case fatality rate was 0.14%.

### Mathematical modelling predictions for the epidemic

Figures 4A-4D show, respectively, the model predictions versus time for SARS-CoV-2 incidence, active-infection prevalence, PCR positivity prevalence (that is also accounting for the prolonged PCR positivity duration^20,21^), and attack rate (ever infection) in the total population, assuming continuation of current levels of social and physical distancing for the coming weeks. Incident infections peaked at 22,630 on May 21, active-infection prevalence peaked at 5.7% on May 23, and PCR positivity prevalence peaked at 18.3% on June 3—there was a 10-day time shift between the epidemic peak based on active-infection prevalence and the peak based on PCR positivity prevalence. The attack rate was estimated at 53.5% on July 12, that is a total of 1.50 million persons have been ever infected. Of these, only 103,128 have been laboratory-confirmed—a diagnosis rate of 6.9%.

**Figure 4.**
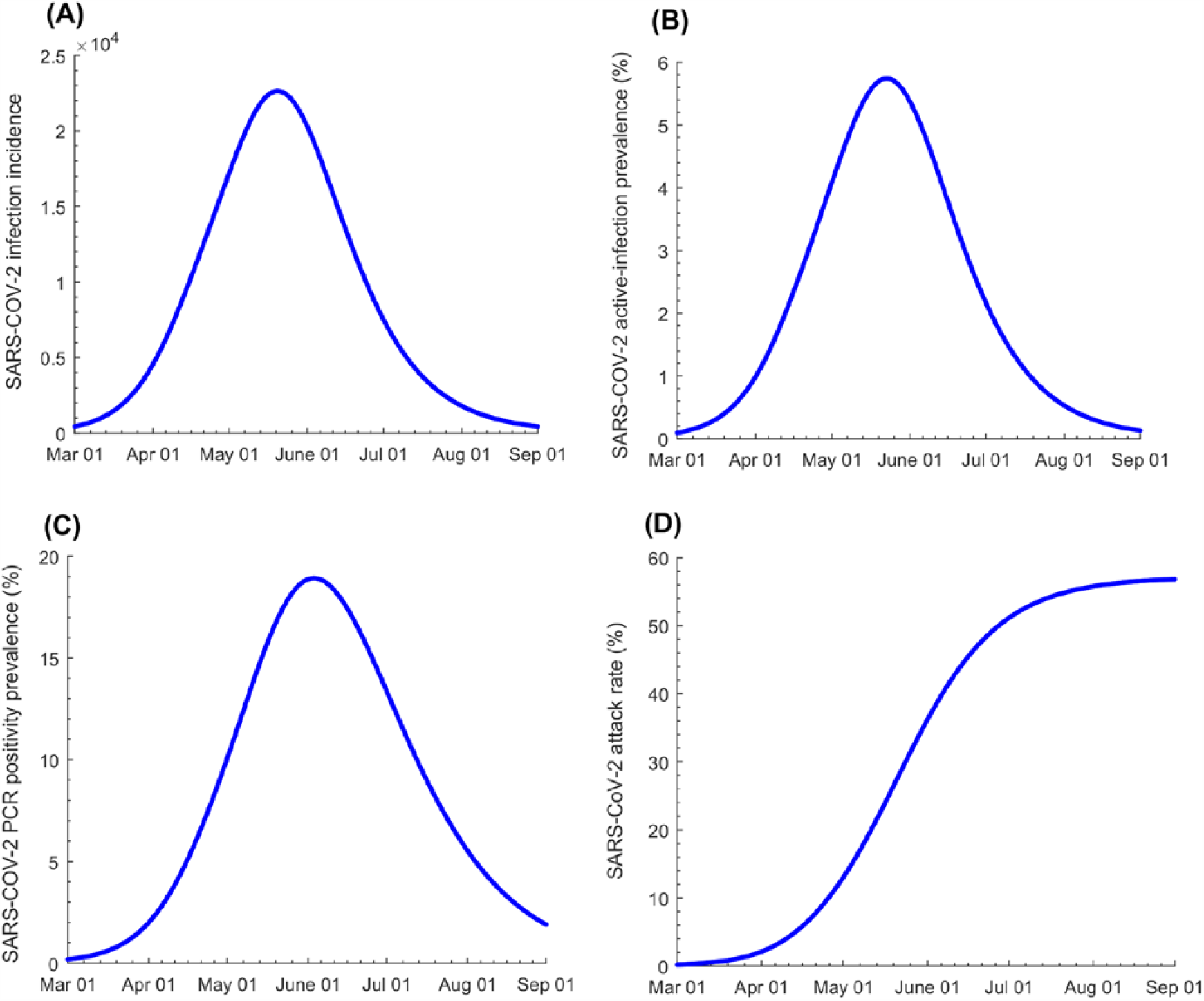
Model predictions for the time evolution of SARS-COV-2 A) incidence, B) active-infection prevalence, C) PCR positivity prevalence, and D) attack rate (ever infection) in the total population of Qatar, assuming continuation of current levels of social and physical distancing for the coming weeks.

The model-estimated *basic* reproduction number (*R_0_*) varied between 1.45-1.68 throughout the epidemic, with the highest values seen in March, during the month of Ramadan (most of May), and after the onset of easing of restrictions in June (Figure S2A). Using a variant of the model incorporating epidemic dynamics by nationality, the model-estimated *R_0_* varied by nationality— it was highest for Nepalese (1.63) and Bangladeshis (1.61) and lowest for Qataris (1.29), with the latter being a proxy for the white collar population of the country who constitutes a fraction of the urban population (Figure S2B). Figure S2C in SM shows the *effective* reproduction number (*R*_*t*_) versus time which declined steadily with the build-up of immunity in the population and was estimated at 0.70 on June 15, setting the day for the onset of easing of the restrictions at the national level.

Figures 3C-3D show the model-predicted *infection* severity rate and *infection* fatality rate, respectively, both calculated with the denominator being the model-estimated cumulative number of *documented and undocumented* infections. By July 5, the infection severity rate (per WHO COVID-19 severity classification)^9^ was predicted to be 0.24 per 100 persons, while the infection fatality rate (per WHO COVID-19 mortality classification)^12^ was predicted to be 0.91 per 10,000 persons.

## Discussion

The SARS-CoV-2 epidemic in Qatar was investigated using different lines of epidemiological evidence and study methodologies, utilizing a centralized and standardized national data-capture system and a national response that emphasized broad testing and provision of early, rapid, standardized, and universal healthcare. Strikingly, these independent lines of evidence converged on similar and consistent findings: Qatar has experienced a pervasive but heterogeneous SARS-CoV-2 epidemic, that is already declining rapidly, apparently due to exhaustion of susceptibles. While the overall epidemic at the national level demonstrated classic susceptible-infected-recovered “SIR” dynamics with an *R_0_* of about 1.6, the national epidemic implicitly included two linked and overlapping but different sub-epidemics. The first affected the labor population, which constitutes the majority of Qatar’s population, and grew rapidly before peaking and then starting to decline. The second and slowly growing sub-epidemic affected the urban population, and appears to be plateauing if not declining slowly, with potential for growth if the easing of the social and physical distancing restrictions proceeds too quickly.

Epidemic intensity in Qatar reflected the unique demographic and residential dwelling structure in this country. The most affected subpopulation was that of the majority population of single CMW living in shared housing accommodations, where workers at a given workplace not only work together during the day, but also typically live together in large dormitories where they share rooms, bathrooms, and cafeteria-style meals. In these settings, the pattern of SARS-CoV-2 transmission showed resemblance to that of influenza outbreaks in schools,^23,24^ and more so to boarding schools,^24^ where the options for effective social and physical distancing are reduced. The differences in exposure by nationality reflected the contribution of each nationality to the labor versus urban population. Yet, these differences may also reflect the structure of social networks in Qatar, as social contacts could be higher among groups who share the same culture, language, and/or national background.

Remarkably, while widespread, the infection has been characterized by relatively low case and infection severity and fatality rates (Figure 3), which were not well above those of a severe seasonal influenza epidemic.^25,26^ The young age profile of the population, with only 8.8% being >50 years of age, appears to explain great part of the low severity—88.4% of confirmed infections were among those <50 years of age. The fact that the epidemic was most intense in the young and healthy CMW population, as opposed to the urban population where all elderly reside, contributed also to the low severity. Indeed, analyses indicated a strong role for age in disease severity and mortality (Figure 2), with even higher effect sizes than elsewhere,^27-30^ possibly because of greater accounting of asymptomatic infection in Qatar. The resourced healthcare system, which was well below the health system threshold even at the epidemic peak, may have also contributed to the low mortality. Emphasis on broad testing coupled with proactive early treatment, such as the treatment of >4,000 cases for pneumonia, may have also limited the number of people who went on to require hospitalization or to develop severe or critical disease.

A notable feature of the epidemic, that is also possibly linked to the population’s young demographic structure, is the large proportion of infections that were asymptomatic or with minimal/mild symptoms for infection to be suspected. The proportion of infections diagnosed at a health facility, somewhat of a proxy for symptomatic infection, was only 60.1%—nearly 40% of infections were diagnosed through contact tracing or surveillance testing in workplaces or residential areas. Among those PCR positive in the community survey, 58.5% reported no symptoms within the last two weeks (Table 3). Given a median PCR Ct value of 28.7 among these asymptomatic persons (Table S5), they are also not likely to develop symptoms following the survey date, as the infection was probably in an advanced stage, if not essentially cleared.^22^ Hospital and isolation facility records show that none of these asymptomatically-infected persons were admitted subsequently with severe or critical disease as of July 10, but there is record indicating that two of them (out of 72) did develop subsequently a symptomatic mild infection.

Meanwhile, 20.5% of those PCR negative in the community survey reported at least one symptom, suggesting that among those who were positive *and* symptomatic, some symptoms may not have been related to SARS-CoV-2 infection (Table 3). Notably, presence of one or two symptoms was not predictive of infection, but presence of three or more symptoms was strongly predictive (Table 3). Although several symptoms were associated (and strongly) with infection (Table S4), very few infected persons reported them (<10%), apart from fever which was reported by 29% of infected persons.

## Limitations

This study has limitations. Symptoms in the community survey were based on self-report, thus introducing potential for recall bias. Although the sampling was intended to be probability-based, most participants were recruited through convenience sampling due to poor response rate suggesting potential for selection bias. However, there was no evidence that PCR positivity differed by method of recruitment (Table 2). Moreover, observed PCR prevalence was similar in both the community survey and the ad-hoc testing campaigns conducted in diverse workplaces and residential areas (Table 2 and Table S6). Analyses of predictors of severity and mortality were limited in scope, not accounting for relevant covariates, such as comorbidities. COVID-19 mortality was based on confirmed hospital deaths and out of hospital deaths per WHO classification,^12^ but such mortality registry may not capture excess deaths caused indirectly by this infection, or deaths misclassified for other causes. Study outcomes may be affected by the sensitivity or specificity of the assays used. However, laboratory methods were based on quality commercial platforms, and each diagnostic method was validated in the laboratory before its use. All results, regardless of the laboratory method used, were also consistent with each other, and specificity of the antibody assay, even if not perfect, may not affect the results given the high antibody prevalence. Of note that the specificity of the antibody assay was reported at 99.8%^14^ by the manufacturer and at 100% by a validation study by Public Health England.^31^ Remarkably, among those who were diagnosed PCR positive or negative >3 weeks before being tested for detectable antibodies (the three weeks to allow for antibodies to be detectable^20^), the percent agreement between the PCR outcome and the antibody outcome was 94.4%, affirming the consistency of both the PCR and antibody methods in diagnosing infection.

## Conclusions

In conclusion, the characterized epidemic of Qatar provides inferences about the epidemiology of this infection. First, SARS-CoV-2 is a highly infectious virus with a large *R_0_*, that is considerably larger than that for other typical cold respiratory viruses.^32-34^ Despite the enforced restrictive social and physical distancing measures, that reduced *R_0_* from as much as 3-4^35,36^ to about 1.6, *R_0_* was well above 1 leading to such large epidemic. Second, while age has been recognized as an important factor since the beginning of the pandemic,^27,37,38 28-30^ it appeared to play even a more critical role in the epidemiology in Qatar than estimated thus far. Not only were serious disease and mortality very strongly linked to being >50 years of age, but most infections in younger persons exhibited no or minimal/mild symptoms. Age may have even played a role in the risk of exposure or susceptibility to the infection (Table 5), as suggested earlier.^39,40^ These findings may suggest that the epidemic expansion in nations with young populations may lead to milder disease burden than previously thought.

## Authors’ contributions

LJA co-conceived and co-designed the study, led the statistical and mathematical modelling analyses, and co-wrote the first draft of the article. HC performed the data analyses and co-wrote the first draft of the article. HHA conducted the mathematical modelling analyses. RB co-conceived and co-designed the study and led the community surveys. All authors contributed to data collection and acquisition, database development, discussion and interpretation of the results, and to the writing of the manuscript. All authors have read and approved the final manuscript.

## Data Availability

Data are available at the discretionary of stakeholders.

## Acknowledgement

We would like to thank Her Excellency Dr. Hanan Al Kuwari, the Minister of Public Health, for her vision, guidance, leadership, and support. We also would like to thank Dr. Saad Al Kaabi, Chair of the System Wide Incident Command and Control (SWICC) Committee for the COVID-19 national healthcare response, for his leadership, analytical insights, and for his instrumental role in enacting the data information systems that made these studies possible. We further extend our appreciation to the SWICC Committee and the Scientific Reference and Research Taskforce (SRRT) members for their informative input, scientific technical advice, and enriching discussions. We also would like to thank Professor David Heymann of the London School of Hygiene & Tropical Medicine and former Assistant Director-General for Health Security and Environment at the World Health Organization for valuable comments and insights on an earlier version of this manuscript. We also would like to acknowledge the efforts of the surveillance team at the Ministry of Public Health as well as all personnel involved in the conduct of the community surveys at the Primary Health Care Centers, Qatar University, Ministry of Public Health, and Hamad Medical Corporation. We further would like to thank all persons who agreed to participate in the community surveys and testing campaigns.

## Supplementary Material

### Text S1. SARS-CoV-2 population-based survey questionnaire

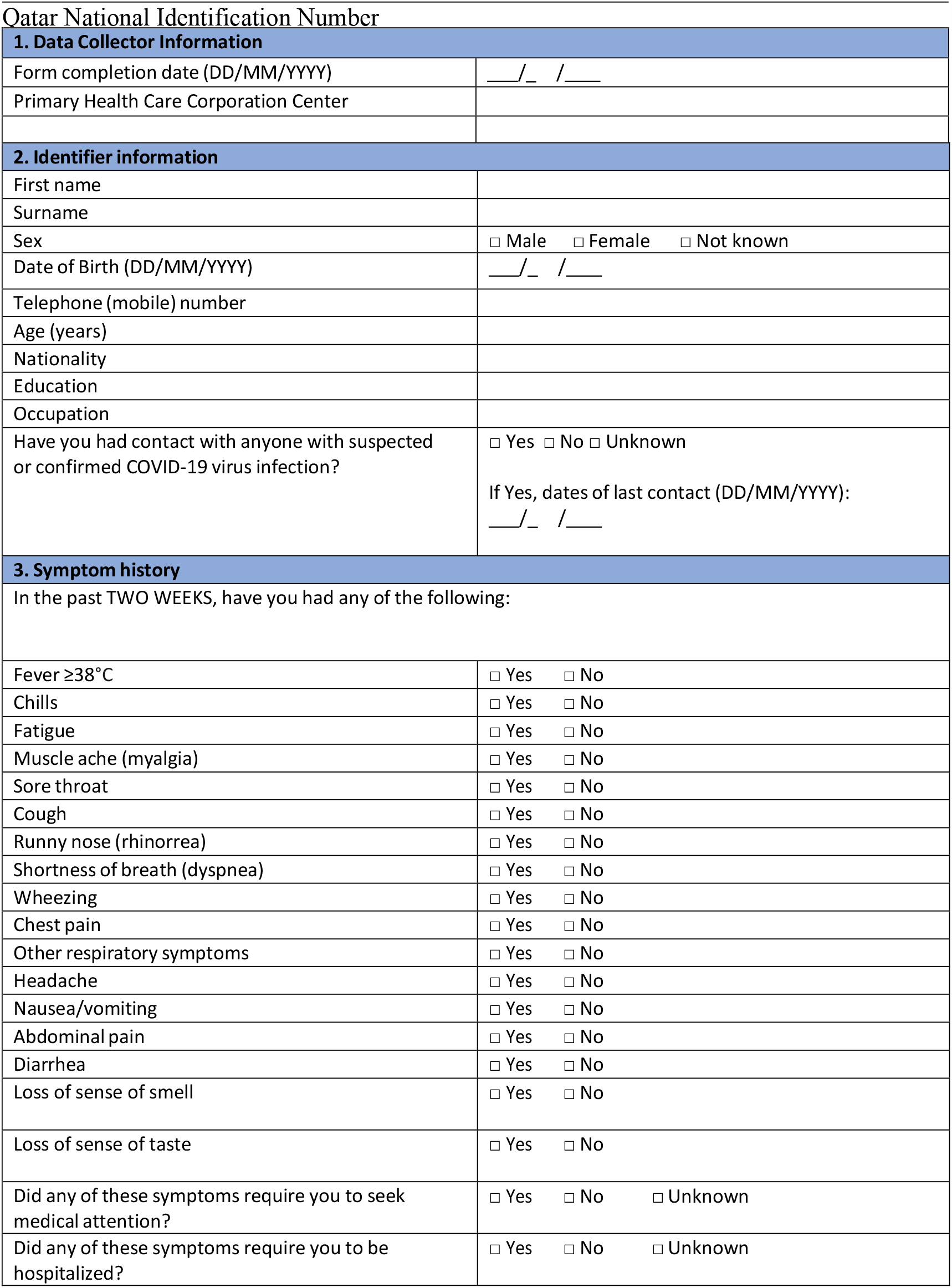

### Text S2. Mathematical model structure and description

We constructed an age-structured deterministic mathematical model to describe the <u>severe acute respiratory syndrome coronavirus 2</u> (SARS-CoV-2) transmission dynamics and disease progression in the population of Qatar (Figure S1). The model stratified the population into compartments according to age group (0-9, 10-19, 20-29,…, ≥80 years), infection status (infected, uninfected), early infection stage (asymptomatic/mild, severe, critical), and disease stage (severe disease or critical disease). This model extends our calibrated mathematical model developed earlier to characterize key attributes of the SARS-CoV-2 epidemic in China.^1^

Epidemic dynamics were described using age-specific sets of coupled nonlinear differential equations. Each age group, *a*, denoted a ten-year age band apart from the last category which grouped together all individuals ≥80 years of age. Qatar’s population size and demographic structure were based on findings of “The Simplified Census of Population, Housing, and Establishments” conducted by Qatar’s Planning and Statistics Authority.^2^ Life expectancy was obtained from the United Nations World Population Prospects database.^3^ All Coronavirus Disease 2019 (COVID-19) mortality was assumed to occur in individuals that are in the critical disease stage, as informed by current understanding of SARS-CoV-2 epidemiology.^4^

**Figure S1.**
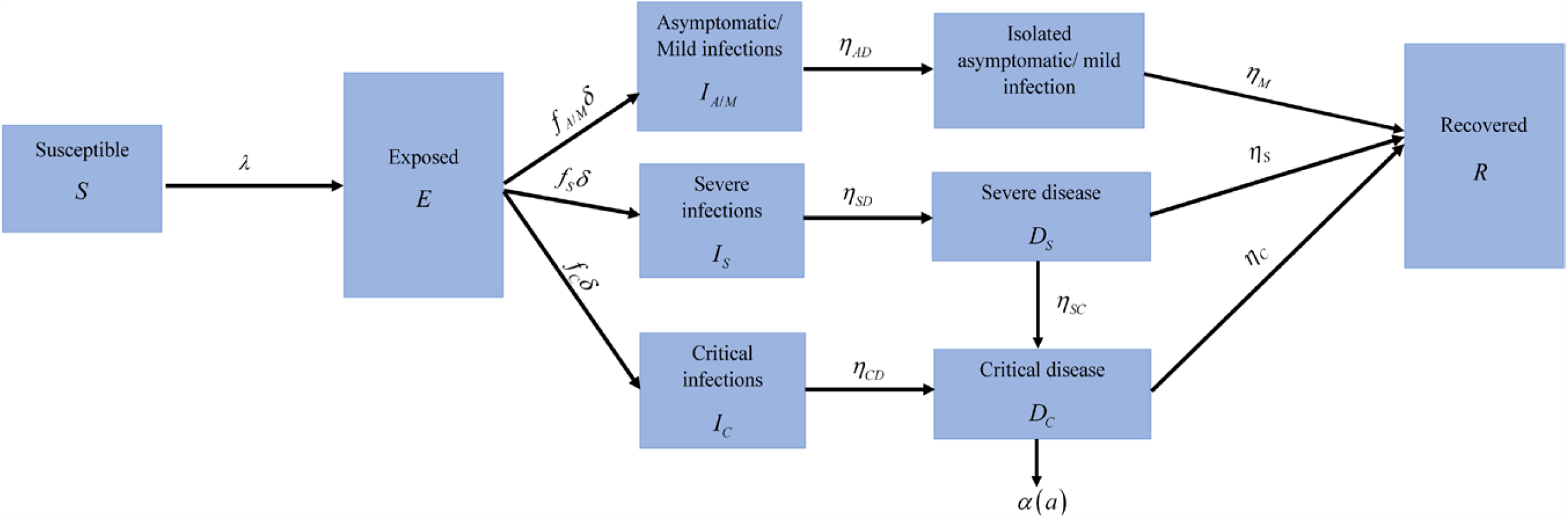
Schematic diagram describing the basic structure of the SARS-CoV-2 mathematical model for Qatar.

The following equations were used to describe the transmission dynamics in the total population:

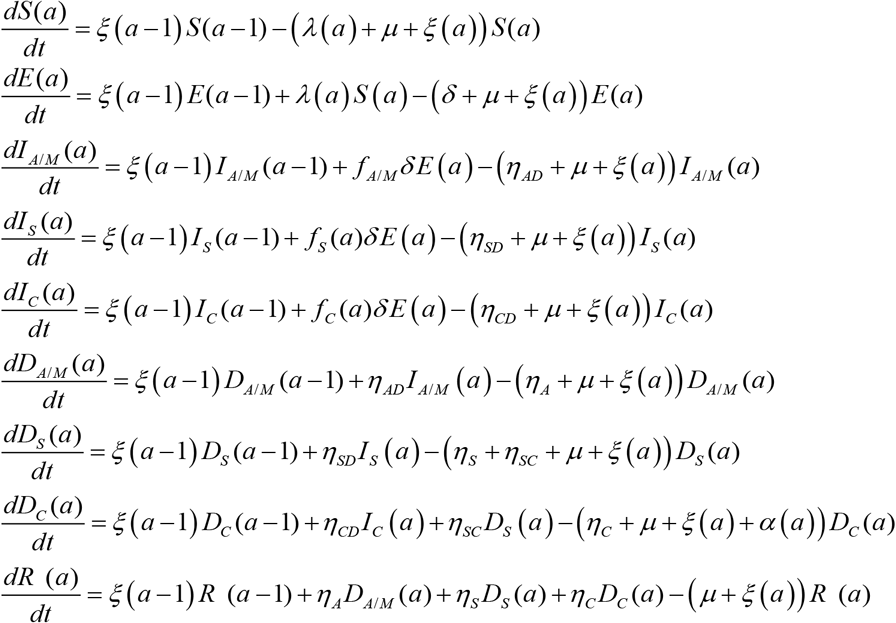

The definitions of population variables and symbols used in the equations are listed in Table S1.

**Table S1.** Definitions of population variables and symbols used in the model

| Symbol | Definition |
| --- | --- |
| $S(a)$ | Susceptible population |
| $E(a)$ | Latently infected population |
| $I_{A/M}(a)$ | Population with asymptomatic/mild infection |
| $I_S(a)$ | Population with severe infection |
| $I_C(a)$ | Population with critical infection |
| $D_{A/M}(a)$ | Population in isolation following diagnosis |
| $D_S(a)$ | Population with hospitalization in acute care beds |
| $D_C(a)$ | Population with hospitalization in intensive care unit care beds |
| $R(a)$ | Recovered population |
| $n_{age}$ | Number of age groups |
| $\xi(a)$ | Transition rate from one age group to the next age group. Here $\xi(0) = \xi(n_{age}) = 0$ |
| $\sigma(a)$ | Susceptibility profile to the infection in each age group |
| $1/\delta$ | Duration of latent infection |
| $\beta$ | Average rate of infectious contacts |
| $1/\eta_{AD}$ | Duration of asymptomatic/mild infection infectiousness |
| $1/\eta_A$ | Duration of asymptomatic/mild infection isolation |
| $1/\eta_{SD}$ | Duration of severe infection infectiousness before isolation and/or hospitalization |
| $1/\eta_S$ | Duration of severe disease following onset of severe disease |
| $1/\eta_{CD}$ | Duration of critical infection infectiousness before isolation and/or hospitalization |
| $1/\eta_C$ | Duration of critical disease following onset of critical disease |
| $1/\eta_{SC}$ | Rate of disease progression from severe disease to critical disease |
| $1/\mu$ | Natural death rate |
| $\alpha(a)$ | Disease mortality rate in each age group |
| $f_{A/M}(a)$ | Proportion of infections that will progress to be asymptomatic/mild infections |
| $f_S(a)$ | Proportion of infections that will progress to be infections that require hospitalization in acute care beds |
| $f_C(a)$ | Proportion of infections that will progress to be infections that require hospitalization in intensive care unit care beds |

The force of infection (hazard rate of infection) experienced by each susceptible population, *S* (*a*), is given by

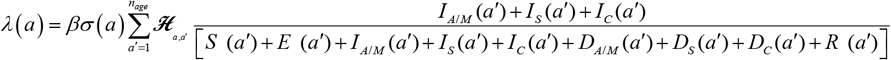

Here, *β* is the average rate of infectious contacts.

The probability that an individual in the *a* age group will mix with an individual in the *a′* age group is determined by an age-mixing matrix, ***ℋ***_*a, a′*_, given by

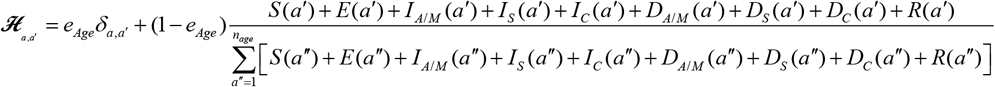

Here, *δ*_*a,a′*_ is the identity matrix and *e*_*Age*_ ∈[0,1] measures the degree of assortativeness in the age mixing. At the extreme *e*_*Age*_ = 0, the mixing is fully proportional, while at the other extreme, *e*_*Age*_ = 1, the mixing is fully assortative, that is individuals mix only with members in their own age group.

### Text S3. Model fitting and parameter values

The model was fitted to data for the 1) time-series of the number of laboratory-confirmed SARS-CoV-2 cases, 2) distribution of laboratory-confirmed cases by age group, 3) time-series of SARS-CoV-2 testing positivity rate, 4) time-series of PCR positivity rate in symptomatic patients with suspected SARS-CoV-2 infection presenting to primary healthcare centers, 5) time-series of new/daily hospital admissions in acute care beds and in ICU care beds, 6) time-series of current hospital occupancy in acute care beds and in ICU care beds, 7) cumulative number of deaths (not time series with the small number of deaths), 8) one community survey for active-infection, and 9) age-distribution of ever infection per the seroprevalence survey.

Model input parameters were based on best available empirical data for SARS-CoV-2 natural history and epidemiology. Model parameter values are listed in Table S2. The following parameters were derived by fitting the model to data: *f* _*A*/ *M*_, *η*_*A*_, *η*_*S*_, *η*_*C*_, and *α*.

**Table S2.** Model assumptions in terms of parameter values

| Parameter | Symbol | Value | Justification |
| --- | --- | --- | --- |
| Duration of latent infection | $1/\delta$ | 3.69 days | Based on existing estimate<sup>5</sup> and based on a median incubation period of 5.1 days<sup>6</sup> adjusted by observed viral load among infected persons<sup>7</sup> and reported transmission before onset of symptoms<sup>8</sup> |
| Duration of infectiousness | $1/\eta_{AD}$;<br>$1/\eta_{SD}$;<br>$1/\eta_{CD}$ | 3.48 days | Based on existing estimate<sup>5</sup> and based on observed time to recovery among persons with mild infection<sup>5,9</sup> and observed viral load in infected persons<sup>7,8,10</sup> |
| Life expectancy in Qatar | $1/\mu$ | 80.7 years | United Nations World Population Prospects database<sup>3</sup> |
| Disease mortality rate in each age group | $\alpha(a)$ | | The distribution and age dependence of COVID-19 mortality was based on the modeled SARS-CoV-2 epidemic in France<sup>11</sup> |
| Age 0-19 years | $RRD1 \times \alpha$ | $RRD1 = 0.1$ | Model-estimated relative risk of death based on the SARS-CoV-2 epidemic in France<sup>11</sup> |
| Age 20-29 years | $RRD2 \times \alpha$ | $RRD2 = 0.4$ | Model-estimated relative risk of death based on the SARS-CoV-2 epidemic in France<sup>11</sup> |
| Age 30-39 years | $\alpha$ | Reference category | Model fitting |
| Age 40-49 years | $RRD3 \times \alpha$ | $RRD3 = 3 \cdot 0$ | Model-estimated relative risk of death based on the SARS-CoV-2 epidemic in France<sup>11</sup> |
| Age 50-59 years | $RRD4 \times \alpha$ | $RRD4 = 10 \cdot 0$ | Model-estimated relative risk of death based on the SARS-CoV-2 epidemic in France<sup>11</sup> |
| Age 60-69 years | $RRD5 \times \alpha$ | $RRD5 = 45 \cdot 0$ | Model-estimated relative risk of death based on the SARS-CoV-2 epidemic in France<sup>11</sup> |
| Age 70-79 years | $RRD6 \times \alpha$ | $RRD6 = 120 \cdot 0$ | Model-estimated relative risk of death based on the SARS-CoV-2 epidemic in France<sup>11</sup> |
| Age 80+ years | $RRD7 \times \alpha$ | $RRD7 = 505 \cdot 0$ | Model-estimated relative risk of death based on the SARS-CoV-2 epidemic in France<sup>11</sup> |
| Proportion of infections that will progress to be infections that require hospitalization in acute care beds | $f_s(a)$ | | The distribution and age dependence of asymptomatic/mild, severe, or critical infections was based on the modeled SARS-CoV-2 epidemic in France<sup>11</sup> |
| Age 0-19 years | $RRS1 \times f_s$ | $RRS1 = 0 \cdot 1$ | Model-estimated relative risk of severe infection based on the SARS-CoV-2 epidemic in France<sup>11</sup> |
| Age 20-29 years | $RRS2 \times f_s$ | $RRS2 = 0 \cdot 5$ | Model-estimated relative risk of severe infection based on the SARS-CoV-2 epidemic in France<sup>11</sup> |
| Age 30-39 years | $f_s$ | Reference category | Model fitting |
| Age 40-49 years | $RRS3 \times f_s$ | $RRS3 = 1 \cdot 2$ | Model-estimated relative risk of severe infection based on the SARS-CoV-2 epidemic in France<sup>11</sup> |
| Age 50-59 years | $RRS4 \times f_s$ | $RRS4 = 2 \cdot 3$ | Model-estimated relative risk of severe infection based on the SARS-CoV-2 epidemic in France<sup>11</sup> |
| Age 60-69 years | $RRS5 \times f_s$ | $RRS5 = 4 \cdot 5$ | Model-estimated relative risk of severe infection based on the SARS-CoV-2 epidemic in France<sup>11</sup> |
| Age 70-79 years | $RRS6 \times f_s$ | $RRS6 = 7 \cdot 8$ | Model-estimated relative risk of severe infection based on the SARS-CoV-2 epidemic in France<sup>11</sup> |
| Age 80+ years | $RRS7 \times f_s$ | $RRS7 = 27 \cdot 6$ | Model-estimated relative risk of severe infection based on the SARS-CoV-2 epidemic in France<sup>11</sup> |
| Proportion of infections that will progress to be infections that require hospitalization in intensive care unit beds | $f_c(a)$ | | The distribution and age dependence of asymptomatic/mild, severe, or critical infections was based on the modeled SARS-CoV-2 epidemic in France<sup>11</sup> |
| Age 0-19 years | $RRC1 \times f_c$ | $RRC1 = 0 \cdot 2$ | Model-estimated relative risk of critical infection based on the SARS-CoV-2 epidemic in France<sup>11</sup> |
| Age 20-29 years | $RRC2 \times f_c$ | $RRC2 = 0 \cdot 3$ | Model-estimated relative risk of critical infection based on the SARS-CoV-2 epidemic in France<sup>11</sup> |
| Age 30-39 years | $f_c$ | Reference category | Model fitting |
| Age 40-49 years | $RRC3 \times f_c$ | $RRC3 = 1.8$ | Model-estimated relative risk of critical infection based on the SARS-CoV-2 epidemic in France<sup>11</sup> |
| Age 50-59 years | $RRC4 \times f_c$ | $RRC4 = 4.7$ | Model-estimated relative risk of critical infection based on the SARS-CoV-2 epidemic in France<sup>11</sup> |
| Age 60-69 years | $RRC5 \times f_c$ | $RRC5 = 10.6$ | Model-estimated relative risk of critical infection based on the SARS-CoV-2 epidemic in France<sup>11</sup> |
| Age 70-79 years | $RRC6 \times f_c$ | $RRC6 = 13.6$ | Model-estimated relative risk of critical infection based on the SARS-CoV-2 epidemic in France<sup>11</sup> |
| Age 80+ years | $RRC7 \times f_c$ | $RRC7 = 8.7$ | Model-estimated relative risk of critical infection based on the SARS-CoV-2 epidemic in France<sup>11</sup> |

**Table S3.** Socio-demographic characteristics of participants in the community survey conducted on May 6-7, 2020 with respect to study recruitment/invitation status

| Socio-demographic characteristics | Invited (probability-based sampling; sample=268) | Not invited (convenience sampling; sample=687) | p-value |
| --- | --- | --- | --- |
|  | N (%<sup>a</sup>) | N (%<sup>a</sup>) |  |
| Sex |  |  |  |
| Males | 216 (80.6) | 559 (81.4) | 0.784 |
| Females | 52 (19.4) | 128 (18.6) |  |
| Age (years) |  |  |  |
| <30 | 36 (13.4) | 184 (26.8) | <0.001 |
| 30-39 | 88 (32.8) | 302 (44.0) |  |
| 40-49 | 64 (23.9) | 125 (18.2) |  |
| 50+ | 80 (29.9) | 76 (11.1) |  |
| Nationality |  |  |  |
| Other<sup>i</sup> | 68 (25.4) | 139 (20.2) | <0.001 |
| Indian | 78 (29.1) | 273 (39.7) |  |
| Filipino | 7 (2.6) | 58 (8.4) |  |
| Sri Lankan | 3 (1.1) | 18 (2.6) |  |
| Egyptian | 30 (11.2) | 52 (7.6) |  |
| Nepalese | 6 (2.2) | 12 (1.8) |  |
| Bangladeshi | 2 (0.8) | 7 (1.0) |  |
| Pakistani | 9 (3.4) | 9 (1.3) |  |
| Sudanese | 6 (2.2) | 10 (1.5) |  |
| Qatari | 59 (22.0) | 109 (15.9) |  |
| Educational attainment |  |  |  |
| Bachelor's degree | 129 (48.1) | 291 (42.4) | 0.001 |
| High school | 90 (33.6) | 282 (41.1) |  |
| Post-graduate studies | 41 (15.3) | 63 (9.2) |  |
| Unknown | 8 (3.0) | 51 (7.4) |  |
| Occupation |  |  |  |
| Healthcare | 15 (5.7) | 30 (4.4) | 0.217 |
| Aviation/Police/Government | 18 (6.8) | 34 (5.0) |  |
| Hospitality/Retail/Education | 33 (12.5) | 66 (9.7) |  |
| Professional/Office | 73 (27.6) | 191 (27.9) |  |
| Transportation/driver | 15 (5.7) | 62 (9.1) |  |
| Construction/Blue collar jobs | 53 (20.0) | 114 (16.7) |  |
| Not employed | 29 (10.9) | 88 (12.9) |  |
| Unknown/Other | 29 (10.9) | 99 (14.5) |  |

<sup>a</sup>Column percentages.

**Table S4.**
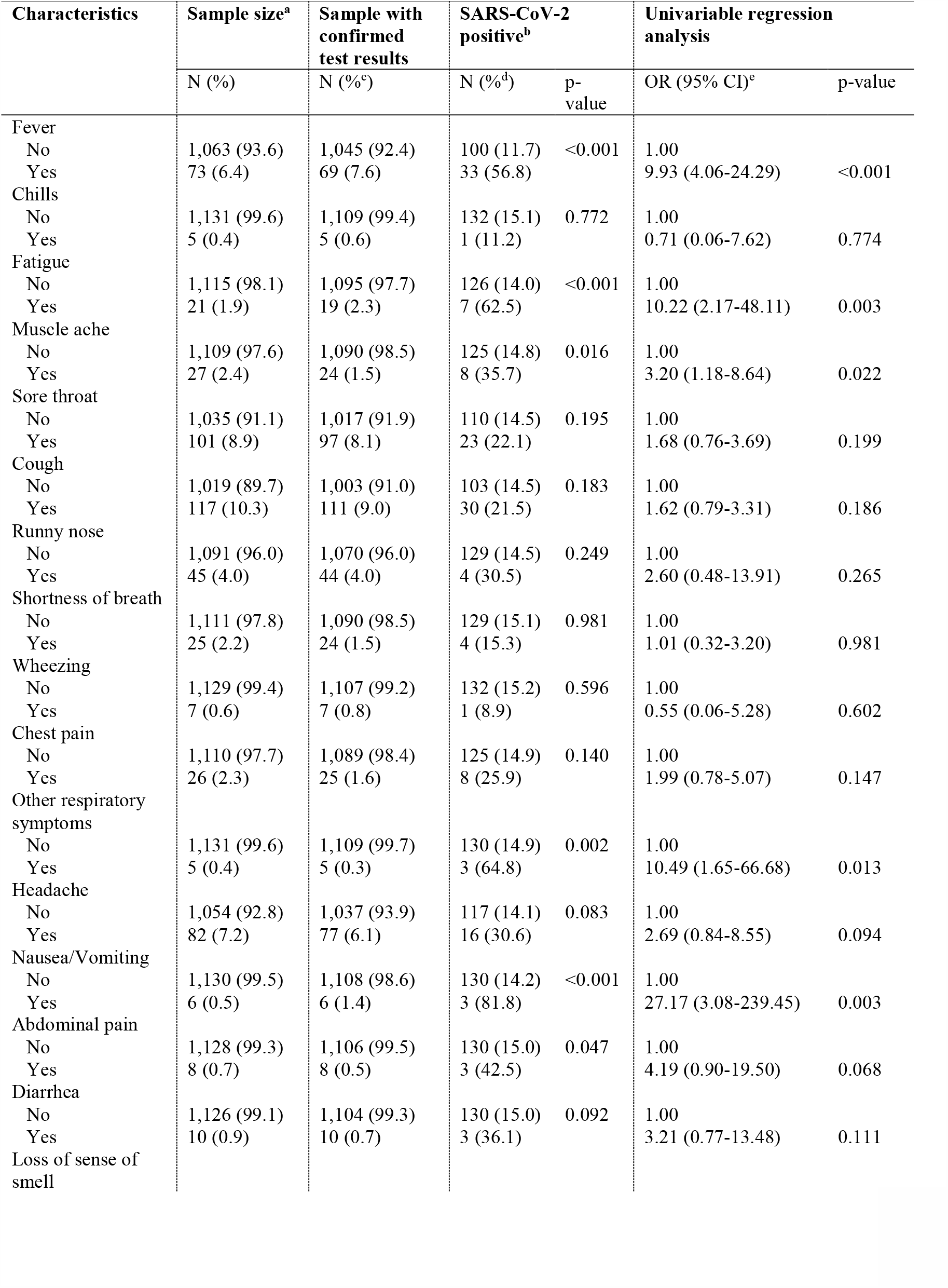
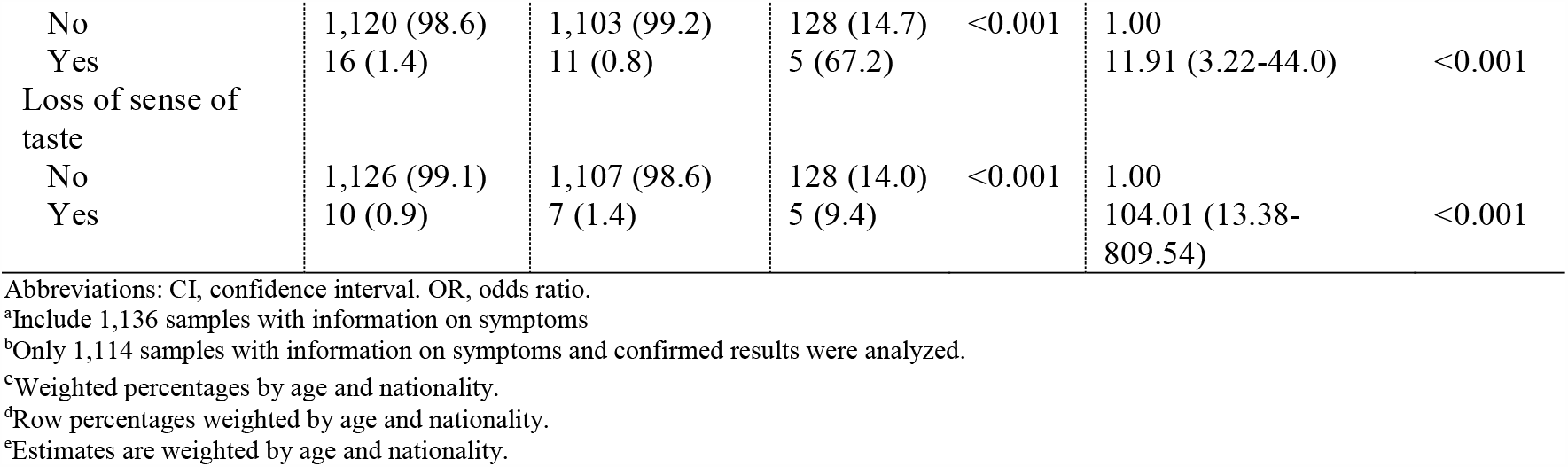
Reported symptoms by individuals tested for SARS-CoV-2 by PCR during the community survey conducted on May 6-7, 2020 and associations with current infection

**Table S5.** Association of PCR Ct values with presence of symptoms for the SARS-CoV-2 confirmed cases identified during the community survey conducted on May 6-7, 2020

| Target protein/gene | Ct value |  |  |  | Ct value <25 |  | Univariable regression analysis |  |
| --- | --- | --- | --- | --- | --- | --- | --- | --- |
|  | N | Mean | Median | Range | N (%<sup>a</sup>) | p-value | OR (95% CI)<sup>a</sup> | p-value |
| N protein<sup>b</sup> |  |  |  |  |  |  |  |  |
| Asymptomatic | 49 | 27.5 | 27.5 | 16.8-35.7 | 21 (36.7) | 0.041 | 1.00 |  |
| Symptomatic | 37 | 21.0 | 21.0 | 15.2-33.0 | 26 (73.3) |  | 4.72 (1.02-21.95) | 0.048 |
| S protein<sup>b</sup> |  |  |  |  |  |  |  |  |
| Asymptomatic | 48 | 26.8 | 28.5 | 16.7-37.0 | 20 (35.8) | 0.037 | 1.00 |  |
| Symptomatic | 37 | 22.5 | 22.4 | 15.4-32.5 | 26 (73.3) |  | 4.92 (1.05-23.08) | 0.044 |
| ORF1ab<sup>b</sup> |  |  |  |  |  |  |  |  |
| Asymptomatic | 48 | 26.1 | 28.0 | 16.4-36.0 | 21 (37.0) | 0.036 | 1.00 |  |
| Symptomatic | 37 | 21.9 | 20.6 | 15.2-33.3 | 27 (74.5) |  | 4.97 (1.05-23.51) | 0.043 |
| E gene<sup>c</sup> |  |  |  |  |  |  |  |  |
| Asymptomatic | 22 | 29.2 | 30.6 | 19.5-36.3 | 6 (20.5) | 0.039 | 1.00 |  |
| Symptomatic | 21 | 26.5 | 26.3 | 17.1-35.8 | 9 (64.7) |  | 7.07 (1.02-49.19) | 0.048 |
| All samples regardless of target protein or gene<sup>d</sup> |  |  |  |  |  |  |  |  |
| Asymptomatic | 71 | 27.2 | 28.7 | 16.8-36.3 | 27 (32.2) | 0.008 | 1.00 |  |
| Symptomatic | 58 | 23.8 | 23.2 | 15.2-35.8 | 58 (69.9) |  | 4.89 (1.47-16.34) | 0.010 |

Abbreviations: CI, confidence interval. OR, odds ratio. PCR, polymerase chain reaction.
<sup>a</sup>Estimates are weighted by age and nationality.
<sup>b</sup>Target assayed using the TaqPath™ COVID-19 Combo Kit (Thermo Fisher Scientific, USA).
<sup>c</sup>Target assayed using the AccuPower SARS-CoV-2 Real-Time RT-PCR Kit (Bioneer, Korea).
<sup>d</sup>Ct values for N protein were used, and complemented by Ct values for E-gene in case of missing values. All individuals with Ct values for S protein or ORF1ab protein also had values for N protein.

**Table S6.** Meta-analysis results for the pooled mean SARS-CoV-2 PCR prevalence in the random testing campaigns conducted in workplaces and in residential areas, up to June 4, 2020

|  | Population groups | Sample |  | SARS-CoV-2 testing positivity (%) |  |  |  | Heterogeneity measures |  |  |
| --- | --- | --- | --- | --- | --- | --- | --- | --- | --- | --- |
|  | Total N<sup>a</sup> | Tested | Positive | Median<sup>a</sup> | Range<sup>a</sup> | Pooled mean | 95% CI | Q<sup>b</sup> (p-value) | I<sup>2c</sup> (% , 95% CI) | Prediction interval<sup>d</sup> (95%) |
| Swab type |  |  |  |  |  |  |  |  |  |  |
| Workplace | 347 | 22,834 | 4,359 | 10.3 | 0.0-100.0 | 15.1 | 13.0-17.3 | 6494.8 (p<0.001) | 94.7 (94.3-95.0) | 0.0-64.2 |
| Residential | 45 | 3,881 | 834 | 19.0 | 0.0-81.5 | 20.3 | 15.4-25.6 | 563.3 (p<0.001) | 92.2 (90.4-93.6) | 0.0-58.0) |
| Testing period |  |  |  |  |  |  |  |  |  |  |
| March<sup>e</sup> | 4 | 552 | 115 | 17.2 | 11.1-36.4 | 17.6 | 9.3-27.7 | 9.7 (p=0.022) | 68.9 (10.2-89.3) | 0.0-62.8 |
| April 4-10 | 32 | 1,856 | 128 | 3.1 | 0.0-70.6 | 4.9 | 1.9-8.9 | 284.5 (p<0.001) | 89.1 (85.7-91.7) | 0.0-35.6 |
| April 11-17 | 28 | 2,476 | 342 | 5.1 | 0.0-59.5 | 10.2 | 4.7-17.2 | 628.0 (p<0.001) | 95.7 (94.6-96.5) | 0.0-58.7 |
| April 18-24 | 56 | 5,422 | 1,130 | 18.0 | 0.0-71.1 | 19.7 | 14.2-25.9 | 1456.2 (p<0.001) | 96.2 (95.6-96.7) | 0.0-70.7 |
| April 25-May 01 | 39 | 2,548 | 568 | 21.6 | 0.0-81.8 | 21.5 | 14.8-29.1 | 659.2 (p<0.001) | 94.2 (92.9-95.3) | 0.0-72.5 |
| May 02-08 | 65 | 4,254 | 838 | 12.1 | 0.0-69.8 | 14.9 | 10.9-19.4 | 849.0 (p<0.001) | 92.5 (91.1-93.6) | 0.0-56.5 |
| May 09-15 | 36 | 3,189 | 837 | 23.0 | 0.0-85.4 | 23.1 | 15.6-31.5 | 909.6 (p<0.001) | 96.2 (95.4-96.8) | 0.0-77.2 |
| May 16-22 | 56 | 3,103 | 829 | 21.8 | 0.0-100.0 | 23.0 | 16.8-29.8 | 892.2 (p<0.001) | 93.8 (92.7-94.8) | 0.0-75.9 |
| May 23-29 | 48 | 2,175 | 362 | 13.4 | 0.0-87.9 | 15.8 | 10.6-21.7 | 471.2 (p<0.001) | 90.0 (87.6-92.0) | 0.0-61.2 |
| May 30-June 04 | 28 | 1,140 | 44 | 1.5 | 0.0-21.7 | 2.5 | 1.0-4.5 | 57.8 (p<0.001) | 53.2 (28.1-69.6) | 0.0-12.7 |
| **Overall** | **392** | **26,715** | **5,193** | **11.5** | **0.0-100.0** | **15.6** | **13.7-17.7** | **7101.2 (p<0.001)** | **94.5 (94.1-94.8)** | **0.0-63.1** |

<sup>a</sup>Testing campaign sample with sample size <10 were grouped together or with the testing campaign sample that has the lowest sample size ≥10
<sup>b</sup>Q: the Cochran's Q statistic is a measure assessing the existence of heterogeneity in effect size (here, SARS-CoV-2 PCR prevalence) across studies.
<sup>c</sup>I<sup>2</sup>: a measure assessing the magnitude of between-study variation that is due to differences in effect size (here, SARS-CoV-2 PCR prevalence) across studies rather than chance.
<sup>d</sup>Prediction interval: a measure estimating the 95% interval of the distribution of true effect sizes (here, SARS-CoV-2 PCR prevalence measures).
<sup>e</sup>Exact dates were not available.

**Table S7.** Basic associations with SARS-CoV-2 severe infection in Qatar based on analysis of the national SARS-CoV-2 PCR testing and hospitalization database. Severe infection was based on WHO classification^12^

| Characteristics | Sample size | SARS-CoV-2 severe infection |  | Univariable analysis |  | Multivariable analysis |  |
| --- | --- | --- | --- | --- | --- | --- | --- |
|  | N (%) | N (%) | p-value | OR (95% CI) | p-value<sup>a</sup> | AOR (95% CI) | p-value<sup>b</sup> |
| Sex |  |  |  |  |  |  |  |
| Females | 13,572 (14.2) | 229 (1.7) | <0.001 | 1.00 |  | 1.00 |  |
| Males | 82,175 (85.8) | 2,207 (2.7) |  | 1.61 (1.40-1.85) | <0.001 | 1.58 (1.35-1.84) | <0.001 |
| Age (years) |  |  |  |  |  |  |  |
| 0-29 | 30,208 (31.5) | 92 (0.3) | <0.001 | 1.00 |  | 1.00 |  |
| 30-39 | 34,487 (36) | 609 (1.8) |  | 5.89 (4.73-7.34) | <0.001 | 5.57 (4.47-6.95) | <0.001 |
| 40-49 | 19,826 (20.7) | 750 (3.8) |  | 12.96 (10.43-16.10) | <0.001 | 12.76 (10.25-15.88) | <0.001 |
| 50-59 | 8,333 (8.7) | 562 (6.9) |  | 24.13 (19.33-30.13) | <0.001 | 25.84 (20.66-32.32) | <0.001 |
| 60-69 | 2,342 (2.4) | 316 (14.1) |  | 53.68 (42.37-68.02) | <0.001 | 59.15 (46.53-75.18) | <0.001 |
| 70-79 | 440 (0.5) | 84 (20.8) |  | 86.16 (62.83-118.14) | <0.001 | 98.24 (70.87-136.17) | <0.001 |
| 80+ | 111 (0.1) | 23 (25.8) |  | 114.02 (68.00-191.18) | <0.001 | 134.99 (78.85-231.11) | <0.001 |
| Nationality |  |  |  |  |  |  |  |
| All other nationalities<sup>c</sup> | 9,501 (9.9) | 239 (2.5) | <0.001 | 1.00 |  | 1.00 |  |
| Indian | 28,111 (29.4) | 450 (1.6) |  | 0.63 (0.54-0.74) | <0.001 | 0.47 (0.39-0.55) | <0.001 |
| Bangladeshi | 13,604 (14.2) | 532 (3.9) |  | 1.58 (1.35-1.85) | <0.001 | 1.42 (1.20-1.68) | <0.001 |
| Nepalese | 17,548 (18.3) | 420 (2.4) |  | 0.95 (0.81-1.11) | 0.517 | 1.06 (0.89-1.26) | 0.531 |
| Pakistani | 5,967 (6.2) | 218 (3.7) |  | 1.48 (1.23-1.78) | <0.001 | 1.13 (0.93-1.38) | 0.209 |
| Sudanese | 1,963 (2.1) | 38 (1.9) |  | 0.76 (0.54-1.08) | 0.129 | 0.57 (0.40-0.81) | 0.002 |
| Sri Lankan | 3,433 (3.6) | 79 (2.3) |  | 0.91 (0.70-1.18) | 0.473 | 0.76 (0.58-0.99) | 0.042 |
| Egyptian | 4,468 (4.7) | 125 (2.8) |  | 1.11 (0.89-1.39) | 0.333 | 0.86 (0.69-1.08) | 0.207 |
| Filipino | 5,116 (5.3) | 207 (4.1) |  | 1.64 (1.36-1.99) | <0.001 | 1.35 (1.11-1.65) | 0.003 |
| Qatari | 6,036 (6.3) | 128 (2.1) |  | 0.84 (0.68-1.04) | 0.112 | 0.82 (0.65-1.04) | 0.101 |
| Testing period |  |  |  |  |  |  |  |
| 05 Feb-31 March | 1,427 (1.5) | 44 (3.1) | <0.001 | 1.00 |  | 1.00 |  |
| 01-15 April | 3,270 (3.4) | 134 (4.2) |  | 1.33 (0.94-1.89) | 0.104 | 1.75 (1.22-2.50) | 0.002 |
| 16-30 April | 9,732 (10.2) | 272 (2.8) |  | 0.89 (0.65-1.23) | 0.494 | 1.26 (0.91-1.77) | 0.169 |
| 01-15 May | 17,221 (18.0) | 483 (2.8) |  | 0.89 (0.65-1.22) | 0.486 | 1.28 (0.93-1.77) | 0.137 |
| 16-31 May | 27,692 (28.9) | 812 (3.0) |  | 0.94 (0.69-1.27) | 0.671 | 1.27 (0.92-1.74) | 0.143 |
| 01-15 June | 23,838 (24.9) | 640 (2.7) |  | 0.85 (0.62-1.16) | 0.311 | 1.17 (0.85-1.61) | 0.348 |
| 16-24 June | 12,567 (13.1) | 51 (0.4) |  | 0.13 (0.08-0.19) | <0.001 | 0.17 (0.11-0.26) | <0.001 |

Abbreviations: AOR, adjusted odds ratio. CI, confidence interval. OR, odds ratio. PCR, polymerase chain reaction.
<sup>a</sup>Covariates with p-value ≤0.1 in the univariable analysis were included in the multivariable analysis.
<sup>b</sup>Covariates with p-value ≤0.05 in the multivariable analysis were considered predictors of SARS-CoV-2 severe infection.
<sup>c</sup>These include all other nationalities residing in Qatar.

**Table S8.**
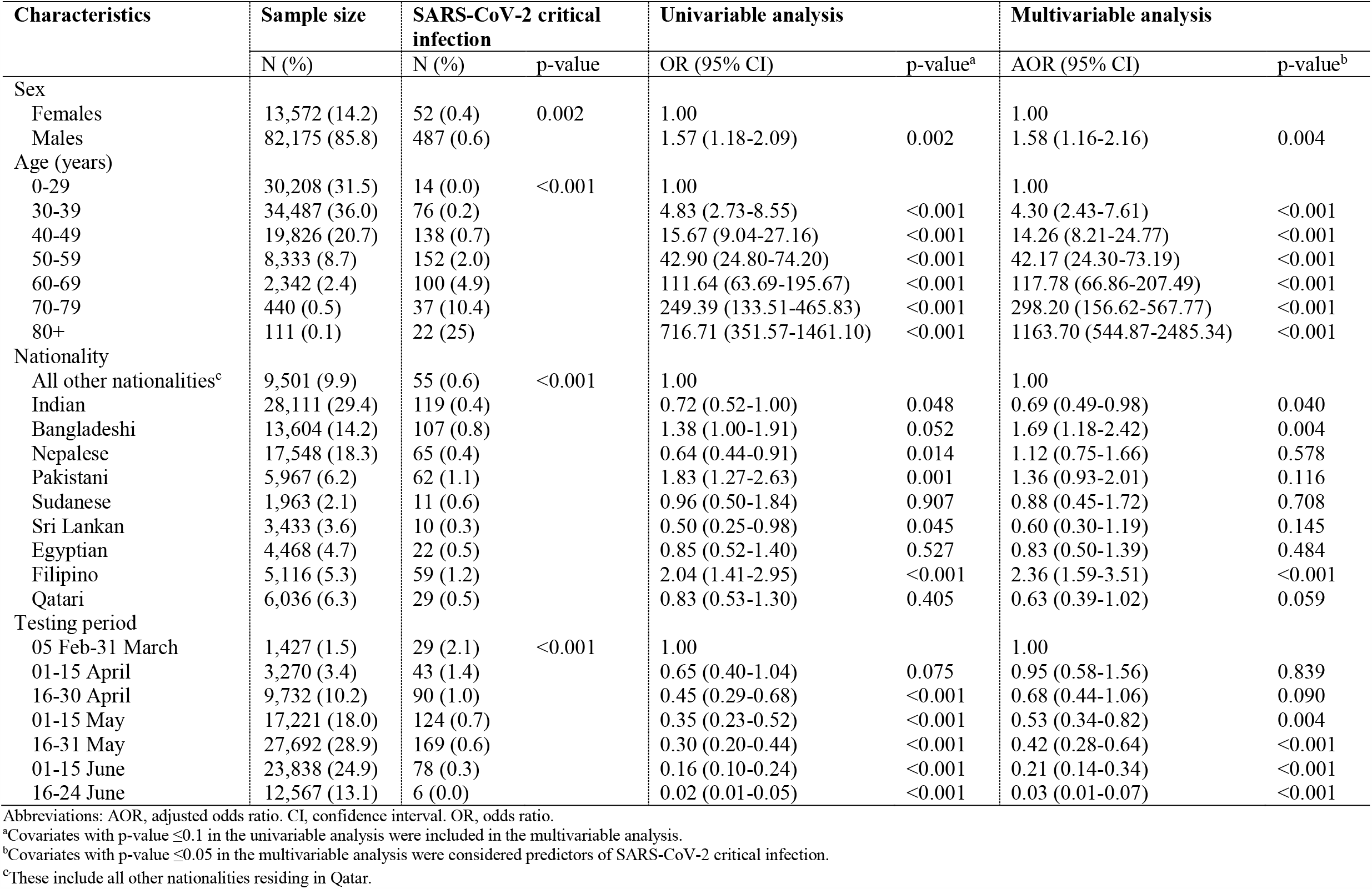
Basic associations with SARS-CoV-2 critical infection in Qatar based on analysis of the national SARS-CoV-2 PCR testing and hospitalization database. Critical infection was based on WHO classification^12^

**Table S9.**
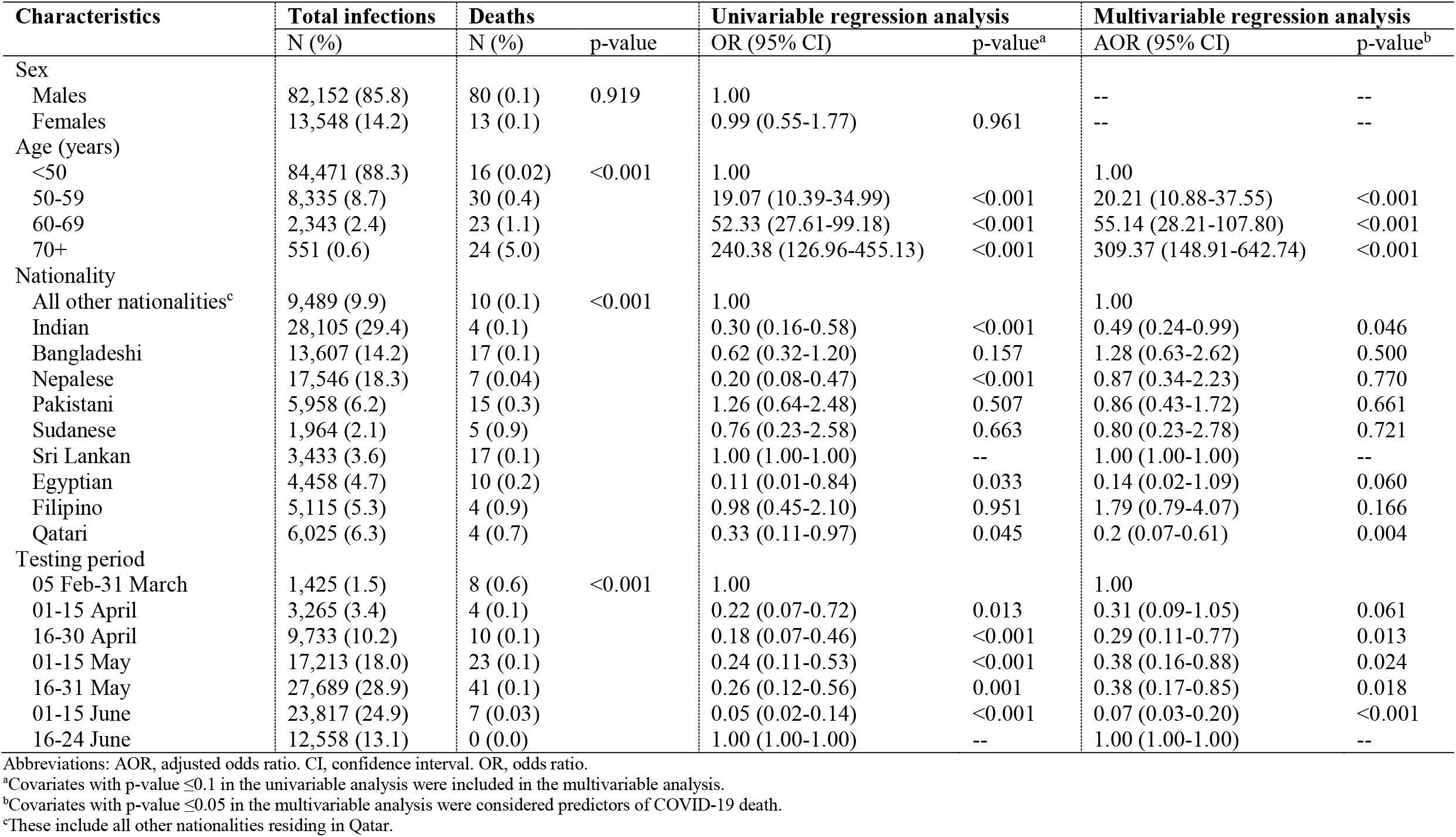
Basic associations with COVID-19 death in Qatar based on analysis of the national COVID-19 mortality database. COVID-19 death was based on WHO classification^13^

**Figure S2.**
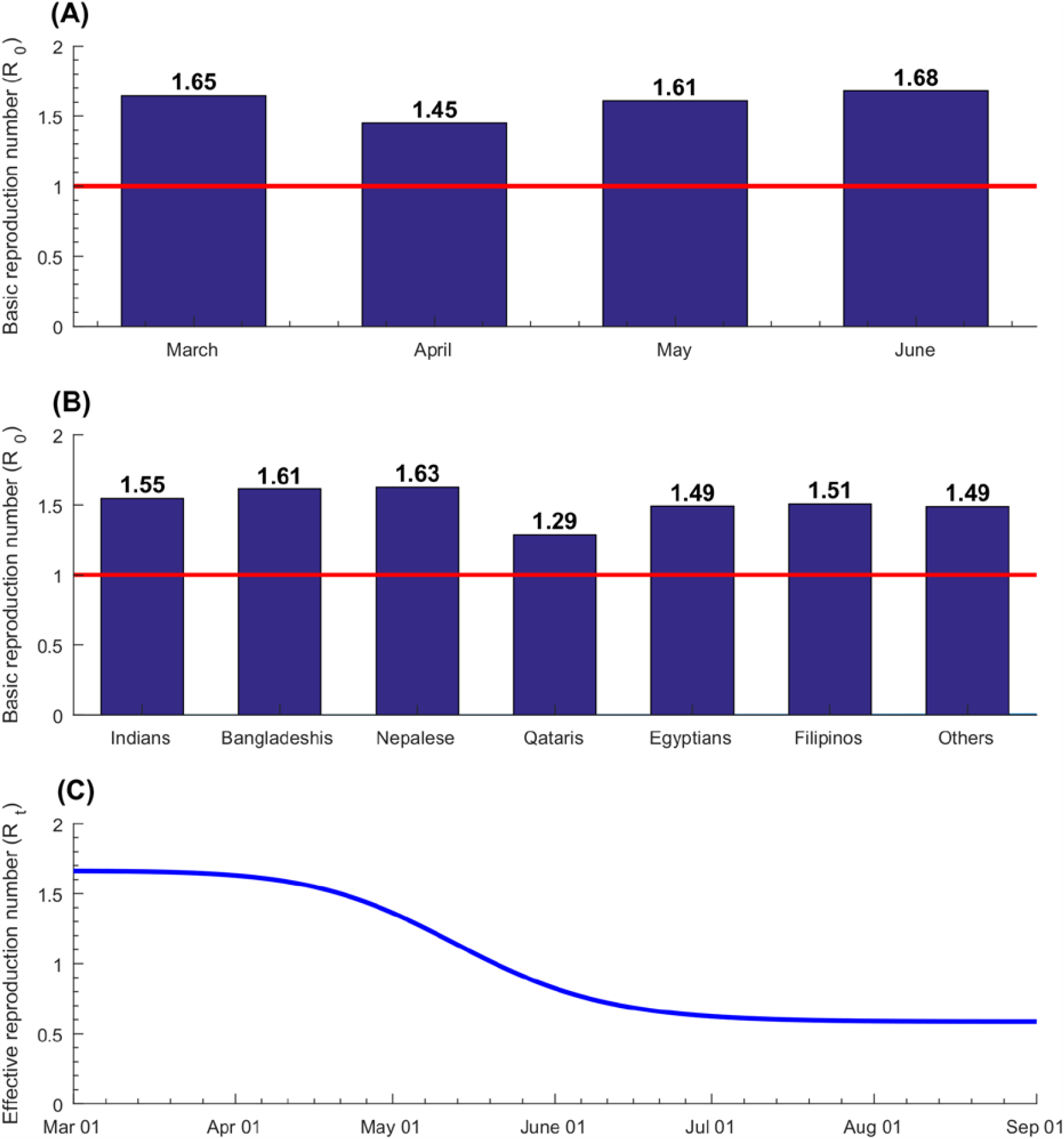
Model predictions for SARS-COV-2 A) basic reproduction number (*R*0 **) over time in the total population of Qatar, B)** *R*_0_ **by nationality, and C) effective reproduction number (** *R*_*t*_ **) in the total population of Qatar**.

